# How radical is radical cure? Site-specific biases in phase-III clinical trials underestimate the effect of radical cure against *Plasmodium vivax* hypnozoites

**DOI:** 10.1101/2021.01.28.21250689

**Authors:** John H. Huber, Cristian Koepfli, Guido España, Narimane Nekkab, Michael T. White, T. Alex Perkins

## Abstract

*Plasmodium vivax* relapses caused by reactivating hypnozoites are a major barrier for elimination and control of this form of malaria. Radical cure is a form of therapy capable of addressing this problem. Recent clinical trials of radical cure have yielded efficacy estimates ranging from 65% to 94%, with substantial variation across trial sites. We performed an analysis of simulated trial data using a transmission model to demonstrate that variation in efficacy estimates across trial sites can arise from differences in the conditions under which trials are conducted. Our analysis revealed that differences in transmission intensity, heterogeneous exposure, and relapse rate can yield efficacy estimates ranging as wide as 12-78%, despite simulating trial data under the uniform assumption that treatment had a 75% chance of clearing hypnozoites. A longer duration of prophylaxis leads to a greater measured efficacy, particularly at higher transmission intensities, making the comparison of the protection of different radical cure treatment regimens against relapse more challenging. We show that vector control and parasite genotyping offer two potential means to yield more standardized efficacy estimates that better reflect protection against relapse. We predict that site-specific biases are likely to contribute to variation in efficacy estimates both within and across phase-III clinical trials. Future clinical trials can reduce site-specific biases by conducting trials in low-transmission settings where reinfections from mosquito biting are less common, by preventing reinfections using vector control measures, or by identifying and excluding likely reinfections that occur during follow-up using parasite genotyping methods.

**AUTHOR SUMMARY:** Radical cure holds promise as a strategy for *Plasmodium vivax* malaria control by clearing the parasites known as hypnozoites that latently infect the liver and cause relapsing infections. The efficacy of radical cure treatment regimens is evaluated in phase-III clinical trials. Recent trial results have noted substantial variation in efficacy estimates across trial sites, complicating the interpretation of the benefit of radical cure. However, *P. vivax* infections identified during the course of the clinical trial could include reinfections from mosquito biting that do not directly reflect the effect of the therapeutic being trialed, potentially biasing efficacy estimates. In this study, we simulated clinical trials to identify the causes and solutions of these site-specific biases. We found that features of both the trial location, such as the transmission intensity, and the trial design, such as the duration of follow-up, lead to an underestimate of the effect of radical cure against hypnozoites. We then demonstrated that vector control and parasite genotyping are two possible strategies to reduce these biases. These insights can be leveraged to aid in the interpretation of past trial results and to help design future clinical trials that minimize site-specific biases.

## INTRODUCTION

*Plasmodium vivax* is the most geographically widespread cause of human malaria, and its burden in 2017 was estimated at 14.3 million clinical cases globally (1). Control of *P. vivax* is challenging due to a unique life stage of the parasite, known as the hypnozoite, which latently infects the livers of individuals with recent *P. vivax* blood-stage infections (2). Hypnozoites activate to cause successive relapsing infections following the initial blood-stage infection, and relapses comprise an estimated 79 – 96% of all *P. vivax* infections (3,4). The prevention of relapses is therefore an ongoing priority for *P. vivax* control (5), and clearance of hypnozoites can be achieved through radical cure treatment with an 8-aminoquinoline, such as primaquine (PQ) or tafenoquine (TFQ).

Recent clinical trials for PQ and TFQ have been conducted in Latin America, sub-Saharan Africa, and Southeast Asia (6–9). The DETECTIVE trial estimated that the recurrence-free efficacies of PQ and TFQ were 74% and 70%, respectively (6), suggesting a large potential impact of radical cure as a first-line *P. vivax* treatment (10). However, each trial noted substantial geographical variation in efficacy estimates. Although potentially reflective of differences in hypnozoite clearance with 8-aminoquinoline treatment among the distinct trial populations (11), the geographical variation in efficacy may have instead been attributable to features of each transmission setting, which would bias the efficacy estimates and limit the interpretability of trial results when evaluating the extent to which radical cure prevents relapse.

The primary endpoint used in recent phase-III clinical trials is freedom from *P. vivax* recurrent infection (6–8), which encompasses relapses caused by hypnozoite activation, reinfections caused by mosquito biting, and recrudescences caused by blood-stage therapeutic failure (12). Only relapses caused by hypnozoite batches (i.e., a group of hypnozoites that derives from one infectious mosquito bite) present prior to treatment directly reflect the extent to which 8-aminoquinoline treatment prevents relapse, or the hypnozoite batch clearance probability (hereafter, clearance probability) (Fig. 1). Because there remains no reliable way to distinguish these relapses from all other recurrent infections using epidemiological data alone, efficacy against recurrent infection is not equal to efficacy against relapse, and the measured efficacy will likely vary depending upon the site in which the trial is conducted. Obtaining standardized estimates of the effect of each 8-aminoquinoline against hypnozoite batches will therefore depend upon the proportion of recurrent infections identified during follow-up that are relapses caused by hypnozoite batches acquired prior to 8-aminoquinoline treatment.

**Fig 1.**
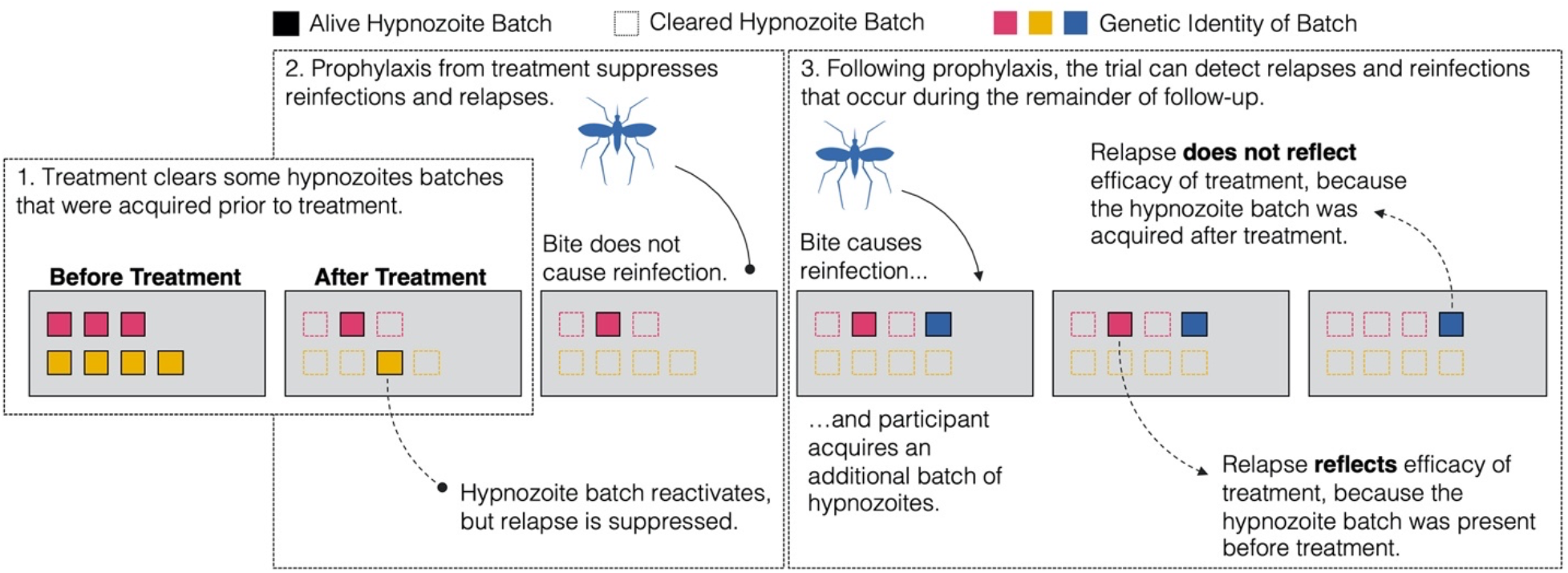
Schematic of potential infection outcomes during trial follow-up. Participants are enrolled in the trial and initially harbor hypnozoite batches (colored squares). The colors represent the genetic identity of each hypnozoite batch. Hypnozoite batches are cleared (unfilled dotted squares) with 8-aminoquinoline treatment according to the per-hypnozoite batch probability of clearance. Treatment with an 8-aminoquinoline provides each participant with a period of prophylaxis during which reinfections (solid line) and relapses (dotted line) are suppressed. Following prophylaxis, trial participants can experience reinfections and relapses, and these reinfections and relapses may be detected during the remainder of follow-up. Only relapses that arise from the activation of hypnozoite batches that were present before 8-aminoquinoline treatment reflect the per-hypnozoite batch clearance probability.

An incongruence between the primary endpoint and the effect that the clinical trial intends to measure has previously been noted as a source of bias in measuring the therapeutic cure rates for *P. falciparum* malaria and tuberculosis (13–15). We propose that the magnitude of this bias could depend upon features of the trial location and the trial design. Although most or all clinical *P. vivax* patients enrolled in clinical trials are expected to carry hypnozoites, the rates at which participants experience reinfections and relapses may vary across trial sites due to various epidemiological features, including transmission intensity, heterogeneous mosquito biting patterns (16), and the relapse phenotype of the *P. vivax* parasite (17). The number of recorded relapses also depends upon the duration of follow-up and the duration of prophylaxis provided by the treatment regimen used in the trial. The extent to which these different features of the trial location and the trial design contribute to bias in efficacy estimates—and impact the ability to measure the clearance probability—has not been explored.

Here, we extend an existing and validated individual-based model of *P. vivax* transmission (10,18) to simulate phase-III clinical trials for radical cure. Individual-based models have been used to design and evaluate clinical trials, both for malaria (19,20) and other infectious diseases (21–24). To maintain consistency with previous trials, we modeled our phase-III clinical trials after the recent DETECTIVE trials for PQ and TFQ (6–8). By varying different features of the trial location and trial design, we sought to understand how site-specific biases in efficacy estimates may arise. We also explored different approaches, such as the use of vector control and parasite genotyping methods, that could be implemented to mitigate site-specific biases and standardize estimates of the effect of radical cure against hypnozoite batches.

## RESULTS

In this analysis, we simulated phase-III clinical trials of an 8-aminoquinoline with an “all-or-none” intervention action and a mean clearance probability of 0.75 across the population. Under this assumption, treatment with the 8-aminoquinoline on average completely cleared hypnozoite batches in 75% of individuals and had no effect on hypnozoite batches in the remaining 25% of individuals. By default, each simulated clinical trial was modeled after the DETECTIVE trials (6–8). Unless otherwise specified, trial participants were followed for 180 days and assessed for clinical and light microscopy (LM)-detectable recurrent infections, and the mean time to first relapse of each *P. vivax* hypnozoite batch was 65 days. Consistent with the DETECTIVE trial (6–8), recurrent infections were left-censored by 32 days to exclude recrudescences caused by blood-stage therapeutic failure, though these infection types were not simulated in the model. A full description of the trial simulations can be found in the Methods and Supplement.

### Effect of Transmission Intensity and Heterogeneity in Biting

We first examined how transmission intensity and the level of heterogeneity in mosquito biting patterns in the trial location interact to affect efficacy estimates. We varied transmission intensity by setting the entomological inoculation rate (EIR) to 1, 10, or 100 infectious bites per person-year and varied the level of heterogeneity in mosquito biting by changing the variance in the distribution of individual-level exposure to mosquito biting.

When exposure to mosquito biting was the same for all trial participants (Fig. 2B), we observed a downward bias in efficacy estimates that increased with transmission intensity (Fig. 2A). For clinical trials of an 8-aminoquinoline with a clearance probability equal to 0.75, light microscopy (LM)-detectable recurrence-free efficacies ranged from 0.70 (IQR: 0.68 – 0.72) at an EIR of 1 to 0.38 (0.32 – 0.43) at an EIR of 100. This downward bias in efficacy occurred due to both frequent reinfection–an infection event that does not reflect the clearance probability–and the reduced detectability of recurrent infections due to higher levels of anti-parasite immunity at higher transmission intensities. As transmission intensity increases, more trial participants were reinfected by mosquitoes, yet parasite densities were reduced and fewer recurrent infections were LM-detectable (Fig. S1, left column), causing the infection profiles of the treatment and control arms to appear more similar and leading to a lower measured efficacy.

**Fig 2.**
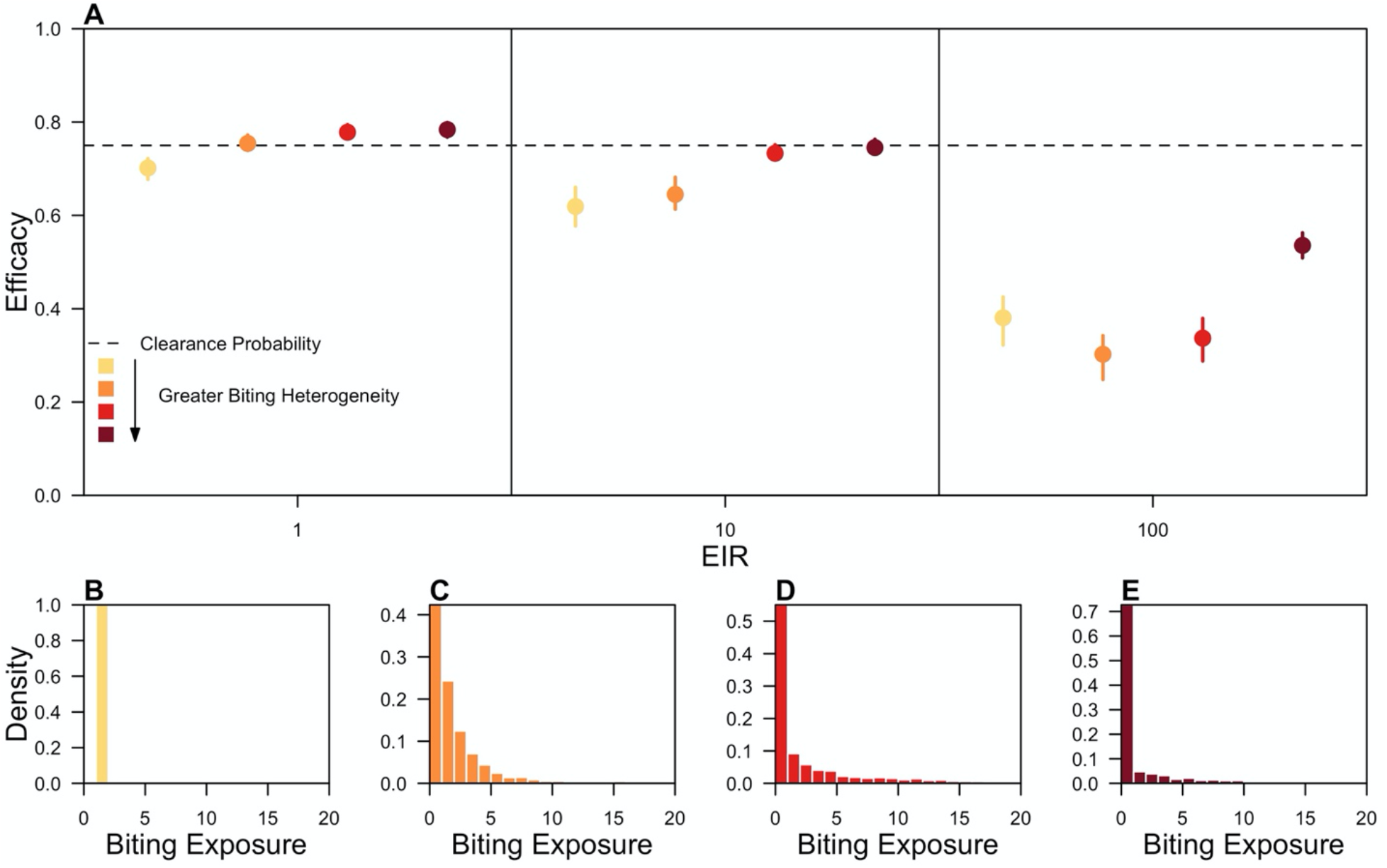
Effect of transmission intensity and heterogeneity in biting on efficacy estimates. (A) LM-detectable recurrence free efficacy estimated from simulated clinical trials is shown at different entomological inoculation rates (EIR) and levels of heterogeneity in biting. Each point represents the median of 200 simulations, and each bar is the interquartile range. The color represents the degree of heterogeneity in individual-level exposure to biting, corresponding to the distributions in (B-E). Darker colors indicate greater heterogeneity in individual-level exposure to biting, and the dotted line is the clearance probability. The distributions of biting propensities are shown from representative simulated trials where the variance in the logged biting propensity was equal to (B) zero, (C) one, (D) two, or (E) three.

At low transmission intensities when reinfection rates were low and heterogeneity in biting was high, our efficacy estimates exceeded the clearance probability, revealing a positive bias. This positive bias occurred because we assumed that radical cure completely cleared hypnozoite batches in a subset of trial participants and did not clear hypnozoite batches in the remainder of trial participants. In trials measuring time to first recurrent infection, those participants that completely clear hypnozoite batches increase the total person-time at risk in the treatment arm, reducing the estimated hazard of recurrent infection in the treatment arm and leading to a competing positive bias that was present in all trial simulations. The magnitude of this bias increases with follow-up time, provided that participants are not reinfected by mosquito biting, and was eliminated if efficacy was instead calculated based upon the proportion at risk using the complete record of recurrent infections (Fig. S2).

In addition to the effect of transmission intensity itself, the effect of heterogeneous biting on efficacy estimates was modulated by the transmission intensity of the trial location. At EIRs of 1 and 10, the downward bias was reduced with greater heterogeneity in biting. Efficacy measured at an EIR of 10 ranged from 0.62 (0.58 – 0.66) under homogeneous biting to 0.75 (0.73 – 0.76) under the highest level of heterogeneity in biting simulated. Heterogeneous biting reduced the downward bias caused by frequent reinfection, because fewer reinfections from mosquito biting occurred during follow-up. As exposure decreased on average with greater heterogeneity in biting, anti-parasite immunity levels were lower, and a greater proportion of recurrent infections were LM-detectable (Fig. S1). The improved detectability of recurrent revealed the differences in the infection profiles across the trial arms, reducing the downward bias due to transmission intensity. At an EIR of 100, the effect of heterogeneous biting on efficacy estimates was non-monotonic (Fig. 2A). Although the percentage of participants reinfected during follow-up decreased monotonically with greater heterogeneity in mosquito biting, the percentage of participants with a detectable reinfection (e.g., clinical or subclinical/LM-detectable) varied non-monotonically and was 36% (35 – 36%), 39% (38 – 40%), 35% (35 – 36%), and 25% (24 – 25%) when the respective variance in exposure to biting was 0, 1, 2, and 3 (Fig. 2B-E).

### Effect of the Rate of Relapse and Duration of Follow-up

We next examined how the rate of relapse of *P. vivax* hypnozoites and the duration of follow-up of trial participants interact to affect efficacy estimates. We considered mean times to relapse of 30, 60, 90, and 180 days and simulated phase-III clinical trials in which participants were followed for 90, 180, 365, or 730 days. To assess the effect of transmission intensity, all trials were simulated at EIRs of 1, 10, and 100, assuming homogeneous biting.

For a given duration of follow-up, efficacy estimates decreased with a longer mean time to first relapse (Fig. 3A). In simulated trials in which participants were followed for 180 days at an EIR of 1, we estimated that the efficacy of an 8-aminoquinoline with a clearance probability of 0.75 decreased from 0.74 (IQR: 0.72 – 0.75) to 0.63 (0.60 – 0.66) as the mean time to first relapse increased from 30 to 180 days. This downward bias occurred because, as the mean time to first relapse increased, fewer trial participants were expected to relapse within the duration of follow-up (Fig. 3B), causing the distribution of times to first recurrent infection to appear more similar across trial arms (Fig. S3, top row). Only 63.2% of participants were expected to relapse by 180 days when the mean time to first relapse was 180 days, compared to 99.8% of participants when the time to first relapse was 30 days.

**Fig 3.**
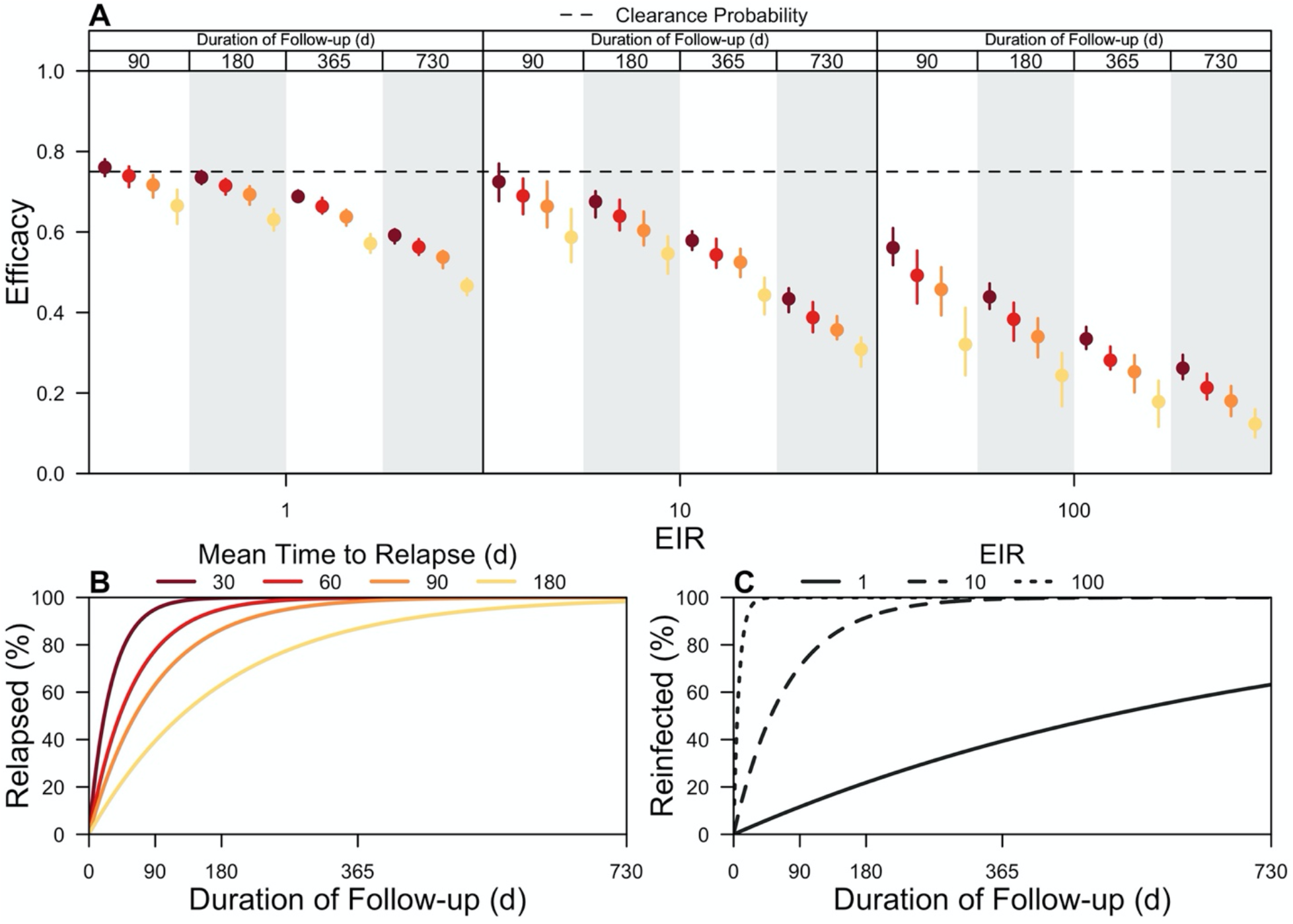
Effect of rate of relapse and duration of follow-up on efficacy estimates. (A) LM-detectable recurrence-free efficacy estimated from simulated clinical trials is shown at different entomological inoculation rates (EIRs), mean times to relapse, and durations of follow-up. Each point is the median of 200 simulations, and each bar is the interquartile range. The color represents the mean time to relapse (30, 60, 90, or 180 days), and each vertical strip for each EIR is the duration of follow-up (90, 180, 365, or 730 days). The dotted line is the clearance probability simulated in each trial. (B) The percentage of trial participants expected to relapse as a function of the duration of follow-up is shown for different mean times to relapse. (C) The percentage of trial participants expected to be reinfected as a function of the duration of follow-up is shown for different EIRs.

For a given mean time to first relapse, our efficacy estimates decreased with a longer duration of follow-up (Fig. 3A). With an EIR equal to 1 and a mean time to first relapse of 30 days, we estimated that the efficacy of an 8-aminoquinoline was 0.76 (0.74 – 0.78) when trial participants were followed for 90 days, compared to 0.59 (0.57 – 0.61) when trial participants were followed for 730 days. Under an alternative scenario in which the EIR was 10, the estimated efficacies were 0.72 (0.68 – 0.77) and 0.43 (0.40 – 0.46) at the respective durations of follow-up. The downward bias occurred because a longer duration of follow-up ensured that more trial participants were reinfected by mosquito biting (Fig. 3C), similarly causing the distribution of times to first recurrent infection to appear more similar across the trial arms. By 90 days of follow-up, 12% and 71% of trial participants were expected to have been reinfected when the respective EIRs were 1 and 10. By 730 days of follow-up, 63% and 100% of trial participants were now expected to have been reinfected at respective EIRs of 1 and 10.

### Effect of Vector Control

Higher transmission intensity biased our efficacy estimates downward, because trial participants in both the treatment and control arms were frequently reinfected (Fig. 2A). We assessed the potential of long-lasting insecticidal nets (LLINs) and indoor residual spraying (IRS) to protect trial participants from reinfection and therefore reduce the downward bias due to transmission intensity. Because the effects of LLINs and IRS depend upon vector bionomics, we varied the absolute proportion of bites that occurred indoors and the absolute proportion of bites that occurred while trial participants were in bed.

Vector control interventions were effective in reducing the downward bias due to transmission intensity in trial locations where the vector was predominantly endophagic (i.e., feeds indoors) (Fig. 4). With an endophagic vector with no preference in biting time and at an EIR of 10, the estimated efficacies improved from 0.58 (0.53 – 0.62) in the absence of vector control to 0.63 (0.58 – 0.67) with LLINs, 0.69 (0.64 – 0.73) with IRS, and 0.70 (0.66 – 0.75) with the combination of LLINs and IRS. By comparison, with an exophagic (i.e., feeds outdoors) vector with no preference in biting time and at an EIR of 10, efficacy estimates improved only slightly from 0.57 (0.52 – 0.61) in the absence of vector control, 0.57 (0.52 – 0.61) with LLINs, and 0.57 (0.52 – 0.61) with IRS to 0.58 (0.55 – 0.63) with the combination of LLINs and IRS (Fig. S4D).

**Fig 4.**
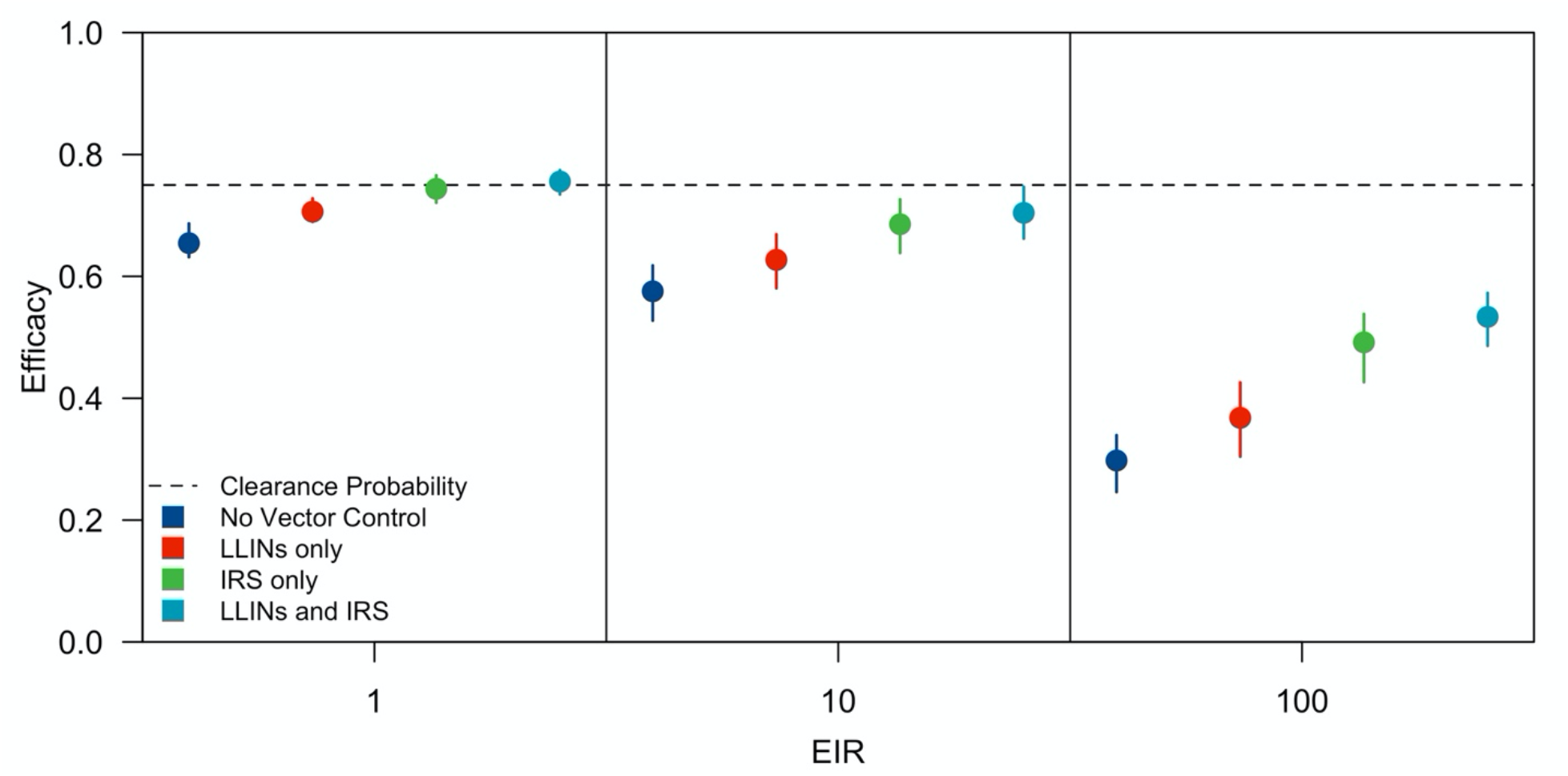
Effect of vector control on efficacy estimates. The impact of LLIN distribution (red), IRS administration (green), and combined LLIN distribution and IRS administration (teal) on LM-detectable recurrence free efficacy estimates is compared to a no-intervention scenario (dark blue) across a range of entomological inoculation rates (EIR). Each point represents the median of 200 simulations, and each bar is the interquartile range. The dotted line is the clearance probability simulated in each trial. The absolute proportion of bites occurring indoors (Φ_I_) was 0.90 and in bed (Φ_B_) was 0.45.

Across all mosquito biting behaviors simulated, the combination of LLINs and IRS was generally more effective than either intervention in isolation in reducing the downward bias in efficacy estimates (Fig. S4). When considering interventions in isolation, IRS was more effective than LLINs in reducing the downward bias, and the distribution of LLINs most improved efficacy estimates under scenarios in which an appreciable number of mosquito bites were taken at night while trial participants were in bed.

### Effect of Parasite Genotyping

Unlike vector control interventions that reduce the downward bias in efficacy by reducing the number of reinfection events among trial participants (Fig. S5), parasite genotyping used in combination with time-to-event data could correct or reduce the downward bias by distinguishing recurrent infection events that directly reflect the clearance probability (i.e., relapses arising from hypnozoite batches acquired prior to treatment) from all other recurrent infections. We assessed the potential of a generic method leveraging genotyping and time-to-event data to correct the bias in efficacy estimates at different transmission intensities and considered how performance characteristics of this method affected the extent to which the bias was corrected by varying its sensitivity and specificity. For the purpose of this simulation study, we defined sensitivity as the probability of correctly identifying relapses caused by hypnozoite batches acquired prior to treatment and specificity as the probability of correctly identifying all other recurrent infections (e.g., reinfections and relapses caused by hypnozoite batches acquired after treatment).

At lower and intermediate transmission intensities (i.e., EIRs of 1 and 10), efficacy estimates changed more with improved sensitivity than improved specificity of the method (Fig. 5). By contrast, at higher transmission intensities, high specificity was needed to reduce the downward bias in efficacy. At an EIR of 100 and assuming 100% sensitivity of the method, the estimated efficacy of an 8-aminoquinoline with clearance probability equal to 0.75 improved from 0.41 (0.37 – 0.47) at 25% specificity to 0.78 (0.73 – 0.81) at 100% specificity. Under the alternative scenario in which specificity was 100%, the estimated efficacy at the respective EIR did not significantly change with improved sensitivity and was 0.78 (0.69 – 0.85) at 25% sensitivity and 0.78 (0.73 – 0.81) at 100% sensitivity. The specificity was more important at higher EIR, because, at an EIR of 100, 100% of participants were expected to be reinfected during follow-up (Fig. 3C). Misclassifying reinfection events as failures of 8-aminoquinoline treatment (i.e., relapses caused by hypnozoites batches acquired prior to treatment) led to a greater downward bias, so a highly specific method was needed in settings where trial participants were frequently reinfected. These effects were less pronounced at low transmission intensities, given that fewer trials participants were reinfected during follow-up.

**Fig 5.**
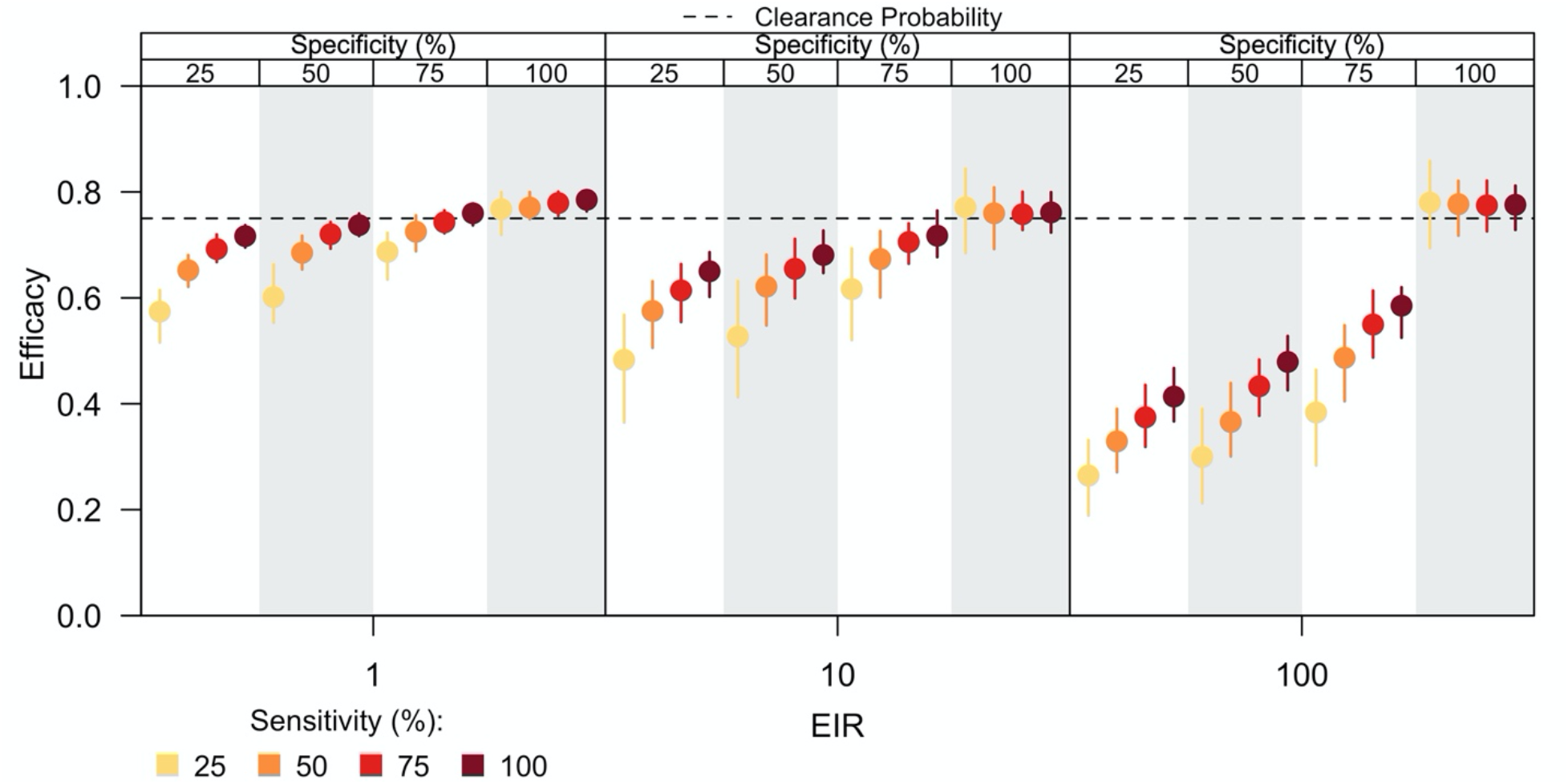
Effect of parasite genotyping on efficacy estimates. The impact of genotyping recurrent infections to estimate the efficacy of radical cure is shown for different sensitivities and specificities of the genotyping method across a range of entomological inoculation rates (EIR). Sensitivity of the genotyping method is the probability of correctly identifying a relapse caused by hypnozoites acquired prior to treatment, and specificity of the genotyping method is the probability of correctly identifying all other recurrent infections. For a given EIR, the vertical panels correspond to the specificity of the genotyping method, and the colors of the points within a given panel denote the sensitivity of the genotyping method. Each point represents the median of 200 simulations, and each bar is the interquartile range. The dotted line is the clearance probability simulated in each trial.

### Effect of the 8-aminoquinoline

The previous analyses considered phase-III trials of an 8-aminoquinoline that provided prophylaxis for 28 days, a duration of time less than the 32-day period of left-censoring used to calculate efficacy. Consequently, prophylaxis did not impact our efficacy estimates. To examine whether a longer duration of prophylaxis biased our efficacy estimates, we simulated phase-III trials for two different 8-aminoquinolines, PQ and TFQ, that provide prophylactic effects for fixed periods of 28 and 45 days, respectively. Because the benefit of prophylaxis increases with transmission intensity, we simulated trials at EIRs equal to 1, 10, and 100, assuming homogeneous biting.

A longer duration of prophylaxis biased our efficacy estimates upward, though the magnitude of the bias depended upon transmission intensity (Fig. 6). In a low-transmission setting (i.e., EIR of 1), our efficacy estimates did not change much with an increased duration of prophylaxis, because a median of 1.3% (1.1 – 1.6%) of participants in the control arm were reinfected between days 32 and 45 post-enrollment—the time period over which TFQ provided prophylactic effects not accounted for by left-censoring. Consequently, at an EIR of 1, we estimated that the efficacies of TFQ and PQ were 0.72 (0.69 – 0.74) and 0.70 (0.68 – 0.72), respectively. At higher EIRs, we estimated a greater efficacy for TFQ than PQ, because participants in the control arm were frequently reinfected whereas participants in the treatment arm were protected from reinfection. On average, the percentage of participants in the control arm who were reinfected between 32- and 45-days post-enrollment was 10% (9.5 – 11%) and 54% (52 – 55%) at EIRs of 10 and 100, respectively. Consequently, we estimated that the respective efficacies of TFQ and PQ were 0.65 (0.62 – 0.69) and 0.63 (0.57 – 0.67) at an EIR of 10 and 0.46 (0.41 – 0.49) and 0.36 (0.32 – 0.41) at an EIR of 100.

**Fig 6.**
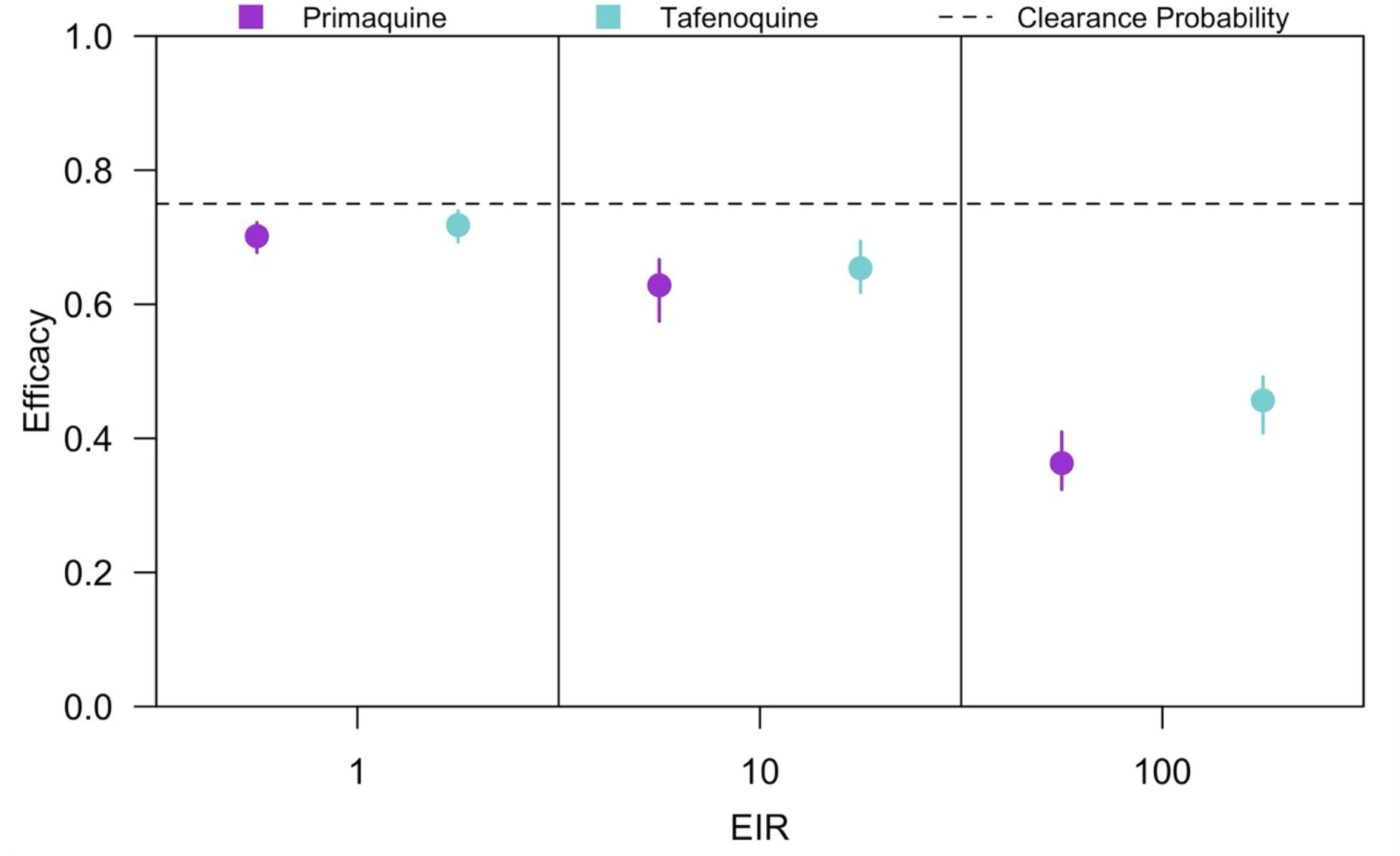
Effect of 8-aminoquinoline on efficacy estimates. LM-detectable recurrence free efficacy estimated from simulated clinical trials is shown for primaquine (purple) and tafenoquine (blue) at different entomological inoculation rates (EIR). Each point is the median of 200 simulations, and each bar is the interquartile range. The dotted line is the clearance probability simulated in each trial.

### Effect of the efficacy metric and infection endpoint

To test whether the efficacy estimates that we obtained were sensitive to the choice of efficacy metric and the infection endpoint, we simulated phase-III clinical trials in which trial participants were assessed for PCR-detectable, LM-detectable, or clinical infections during follow-up. Trials were simulated at EIRs of 1, 10, and 100 assuming homogeneous biting, and efficacy was calculated using three metrics: (1) Cox proportional hazards model, (2) incidence rates, and (3) the proportion at risk.

For all three efficacy metrics considered, increasing transmission intensity caused a downward bias in efficacy estimates, because, at higher transmission intensities, most participants in both the treatment and control arms experienced a reinfection or relapse during follow-up (Fig. 7). For PCR-detectable infections, estimated efficacy was highest if calculated using the Cox proportional hazards model and lowest if calculated using the proportion at risk. This suggested that, in the absence of parasite genotyping, the magnitude of bias caused by reinfection from mosquito biting was more severe when basing efficacy upon incidence rates or the proportion at risk than upon proportional hazards.

**Fig 7.**
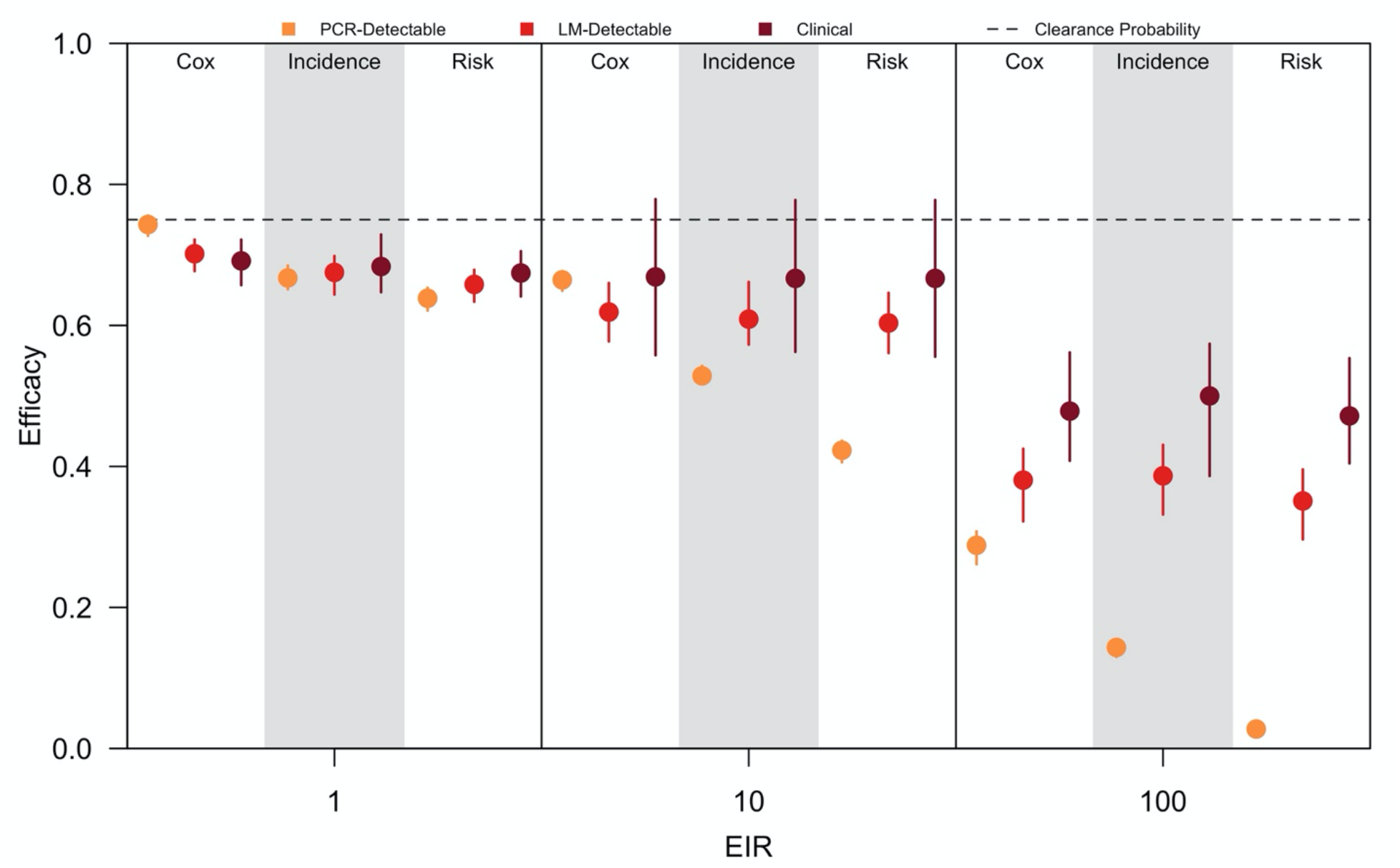
Effect of efficacy metric and infection endpoint on efficacy estimates. Efficacy estimates obtained from simulated clinical trials at different entomological inoculation rates (EIR) is shown when calculated using the Cox proportional hazards model, incidence rates, or the proportion at risk. The infection endpoint was clinical (maroon), LM-detectable (red), or PCR-detectable (orange) recurrent infections identified during follow-up. Each point is the median of 200 simulations, and each bar is the interquartile range. The dotted line is the clearance probability simulated in each trial.

The sensitivity of the assay by which trial participants were assessed for recurrent infections also affected our efficacy estimates, though the direction of the effect depended upon the chosen efficacy metric. For efficacy estimates based upon incidence rates or the proportion at risk, shifting from a more sensitive assay (i.e., PCR-based detection) to a less sensitive assay (i.e., monitoring for clinical infections) reduced the downward bias due to reinfection, particularly at higher transmission intensities. This was due to the fact that the fraction of recurrent infections ascertained increased with a more sensitive assay, causing us to detect more recurrent infections during follow-up and making the incidence rates and proportions at risk appear more similar across the treatment and control arms. By contrast, using the Cox proportional hazards model, the relationship between efficacy estimates and the assay sensitivity depended upon transmission intensity. Across all transmission intensities, clinical infections both occurred less frequently and were detected later than PCR- and LM-detectable infections. Therefore, at an EIR of 1 where reinfection from mosquito biting was less frequent, a more sensitive assay more accurately captured differences in the timing of recurrent infections across trial arms that was attributable to the effect of radical cure. By contrast, at an EIR of 100, nearly all trial participants were reinfected during follow-up. At this transmission intensity, a more sensitive assay instead made the timing of recurrent infections appear more similar across trial arms. Thus, using a less sensitive assay improved efficacy estimates. The non-monotonic relationship observed at the intermediate transmission intensity (i.e., EIR of 10) reflects a change between these two extremes.

## DISCUSSION

Obtaining standardized estimates of the effect of radical cure on *P. vivax* hypnozoites is challenging, because the recurrent infection endpoint used in phase-III clinical trials includes infection events, such as reinfections by mosquito biting, that do not reflect the effect of radical cure on *P. vivax* hypnozoites. In this simulation study, we identified features of the trial setting, including transmission intensity, heterogeneous feeding patterns, and the relapse rate of the *P. vivax* parasite, that affected estimates of efficacy and the utility of clinical trial data to assess the extent to which radical cure prevents relapse (Table 1). We demonstrated that the use of vector control and genotyping methods are two approaches that can reduce and, in some cases, even correct these site-specific biases and yield more standardized estimates of efficacy against relapse.

**Table 1.** Summary of identified biases. A complete description, including the direction and cause, of the biases identified in this analysis are provided.

| Feature of Trial Setting | Direction of Bias | Cause of Bias |
| --- | --- | --- |
| Transmission Intensity (EIR) | Downwards | At higher transmission intensities, there is more frequent reinfection and reduced detectability of recurrent infections. |
| Relapse Rate | Downwards | With a longer time to first relapse, fewer trial participants relapse during follow-up |
| Duration of Follow-up | Downwards | With a longer duration of follow-up, more trial participants are reinfected by mosquito biting. |
| Duration of Prophylaxis | Upwards | With a longer duration of prophylaxis, fewer trial participants are reinfected by mosquito biting. |
| Efficacy Metric | Downwards | Measured efficacy is lower when based upon incidence rates or proportion at risk than upon proportional hazards. |
| Assay Sensitivity | Downward | For incidence rates and the proportion at risk, a more sensitive assay detects more recurrent infections. |
| Intervention Action | Upwards | Efficacy based upon proportional hazards is biased upwards for an “all-or-none” intervention, because the “all” group increases the person-time at risk. |

In the recent GATHER, DETECTIVE, and IMPROV trials (6–9), efficacy estimates varied across trial sites. Interpreting site-specific differences in efficacy is important for past and future clinical trials (25), and our simulation results suggest that the differences in efficacy estimates could be caused by site-specific biases that arise when the infection endpoint used to measure efficacy does not directly reflect the action of the therapeutic being trialed. If the trial sites vary in transmission intensity, our results suggest that efficacy estimates could be lower in high-transmission settings, because trial participants are more frequently reinfected during follow-up, and the detectability of their recurrent infections is reduced. Greater heterogeneity in mosquito biting reduces this downward bias by reducing the number of trial participants who are bitten and possibly reinfected during follow-up, though this effect is non-monotonic at very high transmission intensities due to the predicted interaction with anti-parasite immunity. Trial sites may also differ in the rates at which *P. vivax* hypnozoites relapse (17). A slower rate of relapse (i.e., a longer time to relapse) implies that an appreciable number of trial participants will not yet have relapsed during follow-up, thereby biasing efficacy estimates downward (26). Increasing the duration of follow-up further compounds the bias rather than corrects it, because a long duration of follow-up causes more participants in both the treatment and control arms to become reinfected by mosquito biting, particularly at higher transmission intensities. Finally, frequent reinfection at higher transmission intensities makes the comparison of treatment regimens with different durations of prophylaxis challenging, because a longer duration of prophylaxis prevents more individuals in the treatment arm from reinfection. Ignoring differences in prophylaxis may lead to an overestimate of the effect against hypnozoites for treatment regimens that provide a longer period of prophylaxis, such as tafenoquine co-administered with chloroquine, relative to other treatment regimens that provide a shorter period of prophylaxis, such as primaquine co-administered with chloroquine.

In general, there was greater bias in high-transmission settings, because of the higher risk of recurrent infections being due to reinfections from new mosquito bites. In low-transmission settings, this bias is reduced, although there is the concomitant challenge of recruiting sufficient trial participants. Our simulation results suggest that trial investigators can reduce bias by prospectively preventing reinfections with vector control or by retrospectively accounting for reinfections by leveraging time-to-event and parasite genotyping data.

LLINs and IRS were predicted to be most effective in trial settings with an endophagic vector that takes most bites indoors, so novel vector control interventions, such as spatial repellents, may be needed in settings with an exophagic vector to reduce peri-domestic biting (27). Distribution of LLINs occurred as part of the DETECTIVE trial (6–8), suggesting the feasibility of implementing vector control in a trial context. Entomological data collected prior to trial enrollment could characterize the local vector bionomics and inform the selection of interventions (28,29), and blood meal analysis (30–32) or serological assays (33,34) performed as part of the trial could quantify heterogeneous feeding, a factor that directly affects the magnitude of the site-specific bias.

Our results demonstrate that parasite genotyping could be used in concert with time-to-event data to generate less biased efficacy estimates that better reflect the effect of the 8-aminoquinoline against hypnozoite batches. Although in practice the sensitivity of a genotyping method may be reduced by not observing infections that occurred prior to trial enrollment, the extent to which bias was reduced in our analysis was robust to changes in sensitivity of the method and depended most upon its specificity. High specificity can likely be achieved given the high expected heterozygosity of microsatellite and amplicon deep sequencing panels, as well as the ability to detect minority clones in multiclonal *P. vivax* infections (35–39). Parasites sampled during follow-up were genotyped in the DETECTIVE and IMPROV trials (6–9), and statistical frameworks leveraging genotyping and time-to-event data to distinguish relapses from reinfections have been successfully developed and could be readily integrated into the analysis of past and future clinical trial data (12,39). Nevertheless, this approach may be insufficient to overcome biases introduced by improper trial design, such as a short duration of follow-up relative to the relapse rate in the trial location. Furthermore, higher performance characteristics may be unachievable at higher transmission intensities, where the detectability of recurrent infections is reduced and higher hypnozoite burdens and frequent reinfections limit the information content of genotyping data.

Beyond the factors that make resolving differences in efficacy challenging within the context of a single trial, we identified features of trial design that could limit the comparability of efficacy estimates across trials. Specifically, the details of how the efficacy endpoint was calculated (e.g., time to first infection vs. incidence ratio) and the sensitivity of the assay used to detect recurrent infections resulted in considerable variation in efficacy estimates, particularly at higher transmission intensities. These results suggest that future studies performing meta-analyses of the efficacy of radical cure should consider differences in the trial designs of the clinical trials included.

There are a number of limitations of this analysis. First, we did not calibrate our model to clinical trial data, so our results are not representative of any specific trial settings. However, our simulations encompassed a wide range of epidemiological settings, so the site-specific biases identified in our analysis should reflect variation possible across trial sites. Second, there remains much about hypnozoite biology that is not well understood (40), so our simplified representation of hypnozoite activation and death in our transmission model may not fully capture reality.

Finally, we have an imperfect understanding of the acquisition and loss of anti-parasite and clinical immunity (41). Underestimating the level of anti-parasite and clinical immunity among trial participants may lead to a greater fraction of recurrent infections detected during follow-up than would occur in an actual trial context. This could have led us to overestimate site-specific biases, in which case more refined mathematical representations of immunity could be beneficial for future studies building on ours (10,42).

Our analysis predicted that site-specific biases are likely to occur in phase-III trials for radical cure, and these results suggest that care should be taken in the planning of future trials and the interpretation of trial data. Mathematical modeling that accounts for site-specific biases can aid in the interpretation of clinical trial data and may be useful for identifying future trial sites where biases may be less severe (43). As always, the utility of the insights from mathematical models is improved with additional data, so modeling should be integrated into clinical trial design to identify data needs and inform data collection in order to reduce these biases and improve understanding of radical cure’s potential to control *P. vivax* malaria.

## METHODS

### Ethics Statement

No human subjects were used in this study, so IRB approval was not required.

### Transmission Model

To simulate *Plasmodium vivax* transmission, we used a stochastic, individual-based model developed by White *et al*. (10) and Nekkab *et al*. (18). This model extends the Ross-MacDonald framework of *P. falciparum* transmission to incorporate relapses (44), a characteristic feature of *P. vivax* transmission (45). Calibrated to epidemiological surveys conducted in Papua New Guinea, the Solomon Islands, and Brazil, the model by White *et al*. (10) and Nekkab *et al*. (18) reproduces *P. vivax* transmission dynamics across a range of epidemiological settings.

The rates at which individuals in the population are reinfected and relapse depend upon the respective number of infectious mosquito bites received (i.e., the entomological inoculation rate, or EIR) and the number of hypnozoite batches present in each individual’s liver. The average EIR within the population is determined by the mosquito population dynamics, and individual variation therein is due to heterogeneity in exposure and age-dependent differences in biting. Following successful inoculation with *P. vivax* sporozoites from an infectious mosquito bite, an individual accumulates an additional batch of hypnozoites within the liver and experiences a primary bloodstream infection. In the absence of treatment, each batch of hypnozoites can activate to cause relapses or be cleared naturally from the liver according to fixed rates (Table S1).

The nature of each blood-stage infection (e.g., reinfection or relapse) is determined by the level of anti-parasite and clinical immunity in the infected individual. In the model, the level of anti-parasite immunity determines the probability that a blood-stage infection is detectable by light microscopy (LM) and increases the rate at which low-density infections are cleared. The level of clinical immunity determines the probability that an individual with a blood-stage infection exhibits symptoms.

The levels of anti-parasite and clinical immunity interact to determine the detectability of each blood-stage *P. vivax* infection. The model by White *et al*. (10) and Nekkab *et al*. (18) considers three types of blood-stage infections: (1) sub-microscopic infections detectable by PCR; (2) subclinical infections detectable by LM and PCR; and (3) clinical infections detectable by LM and PCR. Following White *et al*. (10) and Nekkab *et al*. (18), we assumed that each clinical infection was characterized by a fever exceeding 38°C within the last 48 hours and a parasite density greater than 500/µL. Individuals with clinical *P. vivax* infections then seek treatment with anti-malarial drugs according to a probability of treatment-seeking behavior.

Simulation of the *P. vivax* transmission model occurs in two steps. First, the population is initialized at equilibrium according to the analogous set of deterministic compartmental differential equations. Then, the stochastic, individual-based model is simulated with the initialized population. For further description of the model and its assumptions, please refer to the Supplement and the documentation provided in White *et al*. (10) and Nekkab *et al*. (18).

### Trial Overview

We constructed phase-III clinical trials for a generic 8-aminoquinoline (e.g., primaquine or tafenoquine). We simulated trials in order to quantify the recurrence-free efficacy as a measure of the therapeutic effect of radical cure against the hypnozoite batches present in each individual presenting with a clinical *P. vivax* infection confirmed by light microscopy.

We designed our simulated phase-III clinical trials to be comparable to previous trials of PQ and TFQ (6–8). Participants were randomly assigned to the treatment and control arms, and all participants received chloroquine (CQ) upon enrollment to clear the *P. vivax* blood-stage infection. Consistent with the DETECTIVE trial (6,8), participants were followed for 180 days and assessed for asexual parasitemia by light microscopy at days 8, 15, 22, 29, 60, 90, 120, and 180 post-enrollment. For trials simulated with longer duration of follow-up, additional time points were included. See supplement for further details.

### Radical Cure Model

We modeled the action of radical cure in each individual receiving treatment as the per-hypnozoite batch probability of clearance. To allow for heterogeneous action of the intervention (46), we assumed that a proportion *p*_*i*_ of the population belonged to each stratum *i* with per-hypnozoite batch clearance probability, *c*_*i*_. For a population consisting of two strata, the mean per-hypnozoite batch probability of clearance was

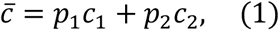

where *p*_2_ = 1 − *p*_1_. This model of heterogeneous action of radical cure generalized the intervention actions commonly considered in the evaluation of clinical trials. Under the “leaky” model (46–48), the action of radical cure is the same across all individuals in the population (*p*_1_ = 1) with 0 ≤ *c*_1_ ≤ 1. Under the “all-or-none” model (46–48), radical cure completely clears hypnozoites in a subset of the population (0 < *p*_1_ < 1; *c*_1_ = 1) and has no effect on the hypnozoites in the remainder of the population (*p*_2_ = 1 − *p*_1_; *c*_2_ = 0). These two models of intervention action could manifest due to host-specific factors, most notably the cytochrome P-450 isoenzyme 2D6 (CYP2D6) genotype, an enzyme involved in metabolizing PQ. For individuals with a low CYP2D6 metabolization phenotype, evidence suggests that PQ may not prevent *P. vivax* relapses (11), though its precise effect on 8-aminoquinoline efficacy remains poorly understood. For example, under an “all-or-none” action, CYP2D6 metabolization could manifest as a binary phenotype with high metabolizers effectively clearing hypnozoite batches and low metabolizers failure to clear hypnozoite batches. Alternatively, under a “leaky” action, all treated individuals partially clear hypnozoite batches. Supplementary analyses revealed that the choice of intervention action did not substantially affect our efficacy estimates generated using the Cox proportional hazards model (Fig. S6). Therefore, we assumed by default that each 8-aminoquinoline had an “all-or-none” action with *p*_1_ = 0.75 and *c*_1_ = 1.

Under the model of heterogeneous action of radical cure, the number of batches of hypnozoites following treatment was distributed as

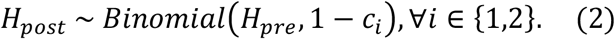

In eq. (2), *H*_*pre*_ is the number of batches of hypnozoite present in the liver prior to treatment, and *c*_*i*_ is the per-hypnozoite batch clearance probability for an individual in stratum *i*.

### Trial Design

We constructed our simulated phase-III clinical trials for radical cure to match the trial design used in past trials for PQ and TFQ (6–8). Accordingly, we considered four phases of the randomized control trial design: (1) recruitment, (2) treatment, (3) vector control, and (3) follow-up.

#### Recruitment

We enrolled individuals in the simulated clinical trial if they presented at a health clinic with febrile illness and were diagnosed by light microscopy with a *P. vivax* infection. Consistent with the DETECTIVE trial (6,8), participants were enrolled if they were at least 16 years of age and had a measured glucose-6-phosphate dehydrogenase (G6DP) activity greater than 70% of the median value of the trial location. Following Nekkab *et al*. (18), we assumed that G6PD activity was measured using the SD Biosensor STANDARD G6PD test (49). This test qualitatively classifies G6PD activity as normal (>70% activity), intermediate (30-70% activity), and low (<30% activity).

Individuals that met the criteria for enrollment were then randomly assigned to the treatment or control arm of the clinical trial. Allocation of participants to each trial arm occurred with equal probability, provided that the current number of participants in each arm was less than the desired sample size of 1,000.

#### Treatment

Upon enrollment, all trial participants were treated with a 3-day course of CQ to clear the blood-stage, asexual parasites (6,8). We assumed that treatment with CQ was 100% effective and that there were no recrudescences among trial participants. Participants in the treatment arm were also treated with an 8-aminoquinoline to clear the hypnozoite reservoir. For individuals in the treatment arm, the number of hypnozoite batches following radical cure was calculated according to eq. (2). We further assumed that treatment with the 8-aminoquinoline provided both blood-stage and liver-stage prophylaxis. Consistent with White *et al*. (10) and Nekkab *et al*. (18), we assumed that the duration of prophylaxis was 28 days for PQ when co-administered with CQ and 45 days for TFQ when co-administered with CQ.

#### Vector Control

As in the DETECTIVE trial (6,8), all participants were provided with a long-lasting insecticidal net (LLIN) to prevent reinfection from mosquitoes during follow-up and therefore decrease the potential bias that may arise in calculating efficacy based on a recurrent infection endpoint. Following Griffin *et al*. (42) and White *et al*. (10), we assumed that vector control measures decrease the probabilities of mosquito biting and successful feeding and increase the probabilities of repellency and death. The magnitude of the effect depends upon the proportion of mosquito bites that occur while individuals are indoors (Φ_*I*_) and in bed (Φ_*B*_), the duration of usage of the LLIN, and the duration of insecticidal activity. We assumed that the probability of usage decayed exponentially over time with a half-life of 3 years. We further assumed that insecticidal activity decayed exponentially over time with a half-life of 2.5 years (18).

We also considered whether the use of indoor residual spraying (IRS) in each participant’s house administered in isolation or in combination with LLINs decreased the potential bias in our efficacy estimates. As with LLINs, we assumed that the use of IRS decreases the probabilities of mosquito biting and successful feeding and increases the probabilities of repellency and death (10,42). To account for the waning effect of IRS over time, we assumed that the insecticidal activity decayed exponentially with a half-life of 6 months (18).

#### Follow-up

We followed each participant for 180 days following enrollment in the clinical trial and treatment with anti-malarial drugs. Consistent with the DETECTIVE trial (6,8), for each participant, we recorded the date of each clinical *P. vivax* infection that occurred within the duration of follow-up. Furthermore, we tested each participant for *P. vivax* asexual parasites using light microscopy on days 8, 15, 22, 29, 60, 90, 120, 160, and 180 post-enrollment. We assumed no mortality among trial participants during the follow-up period.

To examine potential biases that arise from trial surveillance methods, we kept a complete record of all recurrent infections that occurred within the duration of follow-up, including those that would not have been detected under the trial protocol. For each recurrent infection, we recorded the cause (i.e., reinfection or relapse) and the type of blood-stage infection (e.g., sub-microscopic, sub-clinical, or clinical). We further distinguished between relapses caused by hypnozoite batches acquired prior to treatment and relapses caused by hypnozoites batches acquired following treatment. Only relapses caused by hypnozoite batches acquired prior to treatment reflect the action of 8-aminoquinoline treatment against hypnozoites batches.

### Statistical Analyses

We calculated the efficacy of the 8-aminoquinoline used in each clinical trial from the output collected in each respective simulation. We calculated efficacy using multiple metrics in order to examine how different data collected during trial follow-up resolved biases in the efficacy estimates.

Consistent with previous clinical trials (6,8), we used freedom from LM-detectable recurrent infection as the default efficacy metric in our simulation studies. We calculated efficacy using the Cox proportional hazards model (50), which computes the hazard ratio between the treatment and control arms based on the times to first LM-detectable recurrent infection. Following the DETECTIVE trial protocol (6,8), recurrent infections that occurred before 32 days post-enrollment were not included. Given that the timing of each recurrent infection was unobserved, all trial participants were interval censored.

We calculated efficacy using incidence rates computed from the recurrent infections that occurred within the duration of follow-up. We calculated efficacy based on incidence rates as

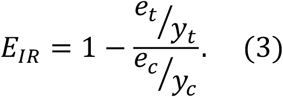

In eq. (3), *e*_*t*_ and *e*_*c*_ are the respective number of infection events in the treatment and control arms, and *y*_*t*_ and *y*_*c*_ are the respective number of person-years of follow-up in the treatment and control arms (48,50).

The magnitude of the measured efficacy also depends upon the action of the radical cure therapeutic. In absence of other sources of bias, we will overestimate efficacy for 8-aminoquinolines that elicit an “all-or-none” response if we calculate efficacy based on incidence rates. This phenomenon is caused by the subset of individuals in the treatment arm who completely clear the hypnozoites from their livers and increase the number of person-years of follow-up (48). To resolve this bias that arises from the assumption of intervention action, we calculated efficacy based on risk as

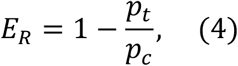

where *p*_*t*_ and *p*_*c*_ are the proportion of individuals in the treatment and control arms that experience a particular infection event within the duration of follow-up.

### Simulation Experiments

We performed simulation experiments to quantify biases in the efficacy estimates that may arise under different transmission settings and other features of the clinical trial design (Table 2). First, we considered how our efficacy estimates varied with transmission intensity and heterogeneity in mosquito biting patterns. Second, we considered how differences in the rate of relapse of the *P. vivax* hypnozoites interact with the duration of the trial follow-up and the transmission intensity to bias our estimates of efficacy. After characterizing these biases, we assessed whether the allocation of vector control measures, such as LLINs and IRS, to trial participants could prevent reinfections and thereby reduce the bias in the efficacy estimates. We next considered whether a method using time-to-event and genotyping data that is capable of distinguishing between different types of *P. vivax* recurrent infections at different sensitivities and specificities could reduce or correct the bias in the efficacy estimates. Then, we measured the extent to which the duration of prophylaxis provided by the treatment regimen biases our estimates of efficacy by preventing reinfections in the treatment arm only. Finally, we considered how the efficacy estimates that we obtained varied with the choice of efficacy metric and infection endpoint.

**Table 2.** Parameters Varied during the Simulation Analyses. The parameters that were varied during each simulation analysis are reported. All other parameters in the transmission model are set to default values and are consistent with the values reported in Tables S1 and S2.

| Variance in Mosquito Biting | Time to Relapse (d) | Follow-up (d) | Prophylaxis (d) |
| --- | --- | --- | --- |
| **Effect of Transmission Intensity and Heterogeneity in Biting** |  |  |  |
| 0, 1, 2, 3 | 65 | 180 | 28 |
| **Effect of Rate of Relapse and Duration of Follow-up** |  |  |  |
| 0 | 30, 60, 90, 180 | 90, 180, 365, 730 | 28 |
| **Effect of Vector Control / Genotyping / Efficacy Metric and Infection Endpoint** |  |  |  |
| 0 | 65 | 180 | 28 |
| **Effect of the 8-aminoquinoline** |  |  |  |
| 0 | 65 | 180 | 28,45 |

For each simulation setting, we simulated a phase-III clinical trial with 1,000 participants in each arm. This sample size was chosen to ensure that the power of our clinical trials exceeded 95% for each transmission intensity and efficacy metric considered (Fig. S7). Each population was simulated for 30 years prior to trial enrollment to ensure that transmission stabilized. Each trial was simulated in a population of 200,000 individuals, and the maximum duration of each trial was 15 years in order to ensure that complete enrollment was attained, even at low transmission intensities. By default, PQ was the 8-aminoquinoline provided to the participants in the treatment arm and when co-administered with CQ provided 28 days of blood-stage and liver-stage prophylaxis during which trial participants could not become reinfected or relapse. The mean per-hypnozoite batch probability of clearance was equal to 0.75. For each simulation setting, we simulated trials in which the therapeutic had an “all-or-none” response. Because the transmission model was stochastic, we ran 200 simulations for each simulation setting and computed the mean and interquartile range for the efficacy estimates across these simulations.

By default, we defined efficacy as freedom from LM-detectable recurrent infection calculated using the Cox proportional hazards model.

### Availability of Code

All code to reproduce the analyses in this study is available on GitHub at https://github.com/johnhhuber/Radical_Cure_Uncertainty.

## Supporting information

Supplemental Methods and Results

## Data Availability

All code to reproduce the analyses in this study is available on Github at https://github.com/johnhhuber/Radical_Cure_Uncertainty.

## Notes

### Competing Interest Statement

The authors have declared no competing interest.

### Funding Statement

JHH acknowledges funding from a National Science Foundation Graduate Research Fellowship and a Richard and Peggy Notebaert Premier Fellowship from the University of Notre Dame. CK received funding from NIH R21-AI137891. NN and MTW were supported by a grant from Medicine for Malaria Venture. NN also received funding from a Pasteur Roux-Cantarini Fellowship. This publication was made possible with partial support from Grant Numbers TL1 TR002531 and UL1TR002529 (A. Shekhar, PI) from the National Institutes of Health, National Center for Advancing Translational Sciences, Clinical and Translational Sciences Award. The funders had no role in study design, data collection and analysis, decision to publish, or preparation of the manuscript.

### Author Declarations

IRB approval was not required for this work, because no human data was used.

