## Supplemental Methods and Results for "How radical is radical cure? Site-specific biases in phase-III clinical trials underestimate the effect of radical cure against *Plasmodium vivax* hypnozoites"

**SUPPORTING INFORMATION**

### Methods

#### *Transmission Model*

We made use of an individual-based model of *Plasmodium vivax* transmission originally developed by White *et al*. (1) and extended by Nekkab *et al*. (2). This model extended the Ross-MacDonald framework of malaria transmission to account for relapses from hypnozoite batches. The individual-based implementation of the model used in this analysis is the stochastic analog of a deterministic compartmental version, and the processes are equivalent across the two models. We detail the compartmental model below and refer the reader to the GitHub repository for details on the implementation of the individual-based model.

*Blood-stage Plasmodium vivax Dynamics in Humans*

In the transmission model, blood-stage *P. vivax* infections are divided into three categories: $I_{PCR}$, $I_{LM}$, and $I_D$. Under this categorization, $I_{PCR}$ are sub-microscopic infections detectable by PCR, $I_{LM}$ are sub-clinical infections detectable by both light microscopy (LM) and PCR, and $I_D$ are clinical infections detectable by both LM and PCR. The model assumes that a fraction $\chi_T$ of clinical infections progress to a treated state $T$ in which treatment with antimalarial drugs rapidly clears the blood-stage parasites. Upon clearing the blood-stage parasites, treated individuals first enter a prophylactic state $P$ in which they cannot be reinfected before ultimately returning to the susceptible state $S$.

The differential equations below detail the processes that govern the movement of individuals between the various compartments. These equations do not yet incorporate relapse, so they are equivalent to the Ross-Macdonald model of *P. falciparum* transmission. Therefore, the force of infection $\lambda_H^0(t - d_E)$ experienced by each individual is driven only by infectious mosquito bites, where $d_E$ is the latent period that accounts for the lag between inoculation of sporozoites and emergence of blood-stage parasites and is set to 10 days.

$$\frac{dS}{dt} = -\lambda_H^0(t - d_E)S + r_{PCR}I_{PCR} + r_P P, \quad (S1)$$

$$\frac{dI_{PCR}}{dt} = -\lambda_H^0(t - d_E)I_{PCR} + \lambda_H^0(t - d_E)(1 - \phi_{LM})(S + I_{PCR}) - r_{PCR}I_{PCR} + r_{LM}I_{LM}, \quad (S2)$$

$$\frac{dI_{LM}}{dt} = \lambda_H^0(t - d_E)I_{LM} + \lambda_H^0(t - d_E)\phi_{LM}(1 - \phi_D)(S + I_{PCR}) + \lambda_H^0(t - d_E)(1 - \phi_D)I_{LM}$$
$$- r_{LM}I_{LM} + r_D I_D, \quad (S3)$$

$$\frac{dI_D}{dt} = \lambda_H^0(t - d_E)\phi_{LM}\phi_D(1 - \chi_T)(S + I_{PCR}) + \lambda_H^0(t - d_E)\phi_D(1 - \chi_T)I_{LM} - r_D I_D, \quad (S4)$$

$$\frac{dT}{dt} = \lambda_H^0(t - d_E)\phi_{LM}\phi_D\chi_T(S + I_{PCR}) + \lambda_H^0(t - d_E)\phi_D\chi_T I_{LM} - r_T T, \quad (S5)$$

$$\frac{dP}{dt} = r_T T - r_P P. \quad (S6)$$

In eqs. (S1-S6), entry into the infection compartments depends not only upon the force of infection $\lambda_H^0(t - d_E)$ but also $\phi_{LM}$, $\phi_D$, and $\chi_T$ which govern the blood-stage infection type and the treatment status. Specifically, $\phi_{LM}$ is the probability that each blood-stage infection is detected by LM, $\phi_D$ is the probability that each LM-detectable blood-stage infection leads to symptoms, and $\chi_T$ is the proportion of clinical infections that progress to the treated state.

*Demography and Age-Dependent Exposure*

Eqs. (S1-S6) do not include changes in population nor the effects of demography on exposure. To account for changes in population, White *et al*. (1) assumed a fixed birth and death rate, $\mu_H$, and a maximum age, $a_{max}$. They then computed the age distribution of the population as a truncated exponential distribution,

$$W(a) = \frac{\mu_H e^{-\mu_H a}}{1 - e^{-\mu_H a_{max}}}. \quad (S7)$$

Furthermore, White *et al*. (1) accounted for age-dependent differences in exposure to mosquito biting. They assumed that the relative exposure with age was described as

$$X(a) = \frac{1}{\omega_{age}}\left(1 - \rho_{age} e^{-\frac{a}{a_0}}\right), \quad (S8)$$

where $\rho_{age}$ is the degree of age-dependent biting and $a_0$ is the reference age of biting. The normalizing constant $\omega_{age}$ was computed as

$$\omega_{age} = 1 - \frac{\rho_{age}\mu_H a_0}{\mu_H a_0 + 1} \frac{1 - e^{-\left(\mu_H + \frac{1}{a_0}\right)a_{max}}}{1 - e^{-\mu_H a_{max}}}. \quad (S9)$$

*Heterogeneity in Biting*

In addition to age-dependent differences in exposure, White *et al.* (1) accounted for heterogeneity in biting that may occur due to other factors, such as attractiveness to mosquitoes, housing quality, and socio-economic status. This heterogeneity in exposure $\zeta$ was assumed to follow a log-Normal distribution,

$$W(\zeta) = \frac{1}{\zeta\sigma\sqrt{2\pi}} e^{-\frac{\log(\zeta)^2}{2\sigma^2}}. \quad (S10)$$

For an EIR equal to $\epsilon$, the standard deviation of individual-level EIRs is equal to $\epsilon\sqrt{e^{\sigma^2} - 1}$, and the force of infection as a function of age and exposure is

$$\lambda_H^0(a,\zeta) = \frac{1}{\omega_{age}}\left(1 - \rho_{age} e^{-\frac{a}{a_0}}\right)\zeta\epsilon. \quad (S11)$$

*Incorporation of Relapses*

To model the transmission dynamics of *P. vivax*, White *et al.* (1) accounted for relapses that are caused by hypnozoites, the dormant hepatic life-stage of the parasite. The model assumed that each infectious mosquito bite leads to the accumulation of a group of hypnozoites, known as a hypnozoite batch. Hypnozoite batches can activate to cause a relapse with rate $f$ and be cleared naturally with rate $\gamma_L$. Thus, if an individual harbors $k$ hypnozoite batches, they will relapse with rate $kf$ and clear one hypnozoite batch with rate $k\gamma_L$. Individuals are assumed to harbor at most $K$ hypnozoite batches.

White *et al.* (1) modeled the proportion $Z_H^k(t,a,\zeta)$ of the population that has $k$ hypnozoite batches as a function of time $t$, age $a$, and exposure $\zeta$. This is captured in the set of partial differential equations:

$$\frac{\partial Z_H^0}{\partial t} + \frac{\partial Z_H^0}{\partial a} + \frac{\partial Z_H^0}{\partial \zeta} = -\lambda_H^0(t - d_E, a - d_E, \zeta) Z_H^0 + \gamma_L Z_H^1 + \mu_H W(\zeta) I(a = 0) - \mu_H Z_H^0, \quad (S12)$$

$$\frac{\partial Z_H^k}{\partial t} + \frac{\partial Z_H^k}{\partial a} + \frac{\partial Z_H^k}{\partial \zeta} = -\lambda_H^0(t - d_E, a - d_E, \zeta) Z_H^k + \lambda_H^0(t - d_E, a - d_E, \zeta) Z_H^{k-1} - \gamma_L k Z_H^k + \gamma_L (k+1) Z_H^{k+1} - \mu_H Z_H^k, \quad (S13)$$

$$\frac{\partial Z_H^K}{\partial t} + \frac{\partial Z_H^K}{\partial a} + \frac{\partial Z_H^K}{\partial \zeta} = \lambda_H^0(t - d_E, a - d_E, \zeta) Z_H^{K-1} - \gamma_L K Z_H^K - \mu_H Z_H^K. \quad (S14)$$

*Acquisition of Immunity*

The model considers two types of immunity: anti-parasite ($A_P$) and clinical ($A_c$). White *et al.* (1) assumed that anti-parasite immunity reduced $\phi_{LM}$ and increased $r_{PCR}$ and that clinical immunity reduced $\phi_D$. Each individual's level of immunity depends upon age and exposure, and additional blood-stage infections boost immunity. Thus, for a given force of infection $\lambda$, the rate of immunity boosting is $\frac{\lambda}{\lambda u+1}$, where *u* is the length of a refractory period for which immunity cannot continue to be boosted. The dynamics of anti-parasite immunity in the population as a function of time *t*, age *a*, and exposure $\zeta$ is captured by the following set of partial differential equations:

$$\frac{\partial A_P^0}{\partial t} + \frac{\partial A_P^0}{\partial a} + \frac{\partial A_P^0}{\partial \zeta} = -\lambda_H^0(t - d_E, a - d_E, \zeta) A_P^0 - r_{par} A_P^0 + \gamma_L A_P^1 - \mu_H A_P^0, \quad (S15)$$

$$\frac{\partial A_P^k}{\partial t} + \frac{\partial A_P^k}{\partial a} + \frac{\partial A_P^k}{\partial \zeta} = \frac{\lambda_H^k(t-d_E, a-d_E, \zeta)}{\lambda_H^k(t-d_E, a-d_E, \zeta)u_{par} + 1} - \lambda_H^k(t-d_E, a-d_E, \zeta)A_P^k + \lambda_H^{k-1}(t-d_E, a-d_E, \zeta)A_P^{k-1} - r_{par}A_P^k - \gamma_L k A_P^k + \gamma_L(k+1)A_P^{k+1} - \mu_H A_P^k, \quad (S16)$$

$$\frac{\partial A_P^K}{\partial t} + \frac{\partial A_P^K}{\partial a} + \frac{\partial A_P^K}{\partial \zeta} = \frac{\lambda_H^K(t-d_E, a-d_E, \zeta)}{\lambda_H^K(t-d_E, a-d_E, \zeta)u_{par} + 1} + \lambda_H^{K-1}(t-d_E, a-d_E, \zeta)A_P^{K-1} - r_{par}A_P^K - \gamma_L K A_P^K - \mu_H A_P^K. \quad (S17)$$

Similarly, the dynamics of clinical immunity in the population as a function of time *t*, age *a*, and exposure $\zeta$ is captured by the following set of partial differential equations:

$$\frac{\partial A_C^0}{\partial t} + \frac{\partial A_C^0}{\partial a} + \frac{\partial A_C^0}{\partial \zeta} = -\lambda_H^0(t-d_E, a-d_E, \zeta)A_C^0 - r_C A_C^0 + \gamma_L A_C^1 - \mu_H A_C^0, \quad (S18)$$

$$\frac{\partial A_C^k}{\partial t} + \frac{\partial A_C^k}{\partial a} + \frac{\partial A_C^k}{\partial \zeta} = \frac{\lambda_H^k(t-d_E, a-d_E, \zeta)}{\lambda_H^k(t-d_E, a-d_E, \zeta)u_C + 1} - \lambda_H^k(t-d_E, a-d_E, \zeta)A_C^k + \lambda_H^{k-1}(t-d_E, a-d_E, \zeta)A_C^{k-1} - r_C A_C^k - \gamma_L k A_C^k + \gamma_L(k+1)A_C^{k+1} - \mu_H A_C^k, \quad (S19)$$

$$\frac{\partial A_C^K}{\partial t}+\frac{\partial A_C^K}{\partial a}+\frac{\partial A_C^K}{\partial \zeta} = \frac{\lambda_H^K(t-d_E,a-d_E,\zeta)}{\lambda_H^K(t-d_E,a-d_E,\zeta)u_C+1}+\lambda_H^{K-1}(t-d_E,a-d_E,\zeta)A_C^{k-1}-r_C A_C^K-\gamma_L K A_C^K - \mu_H A_C^K. \quad (S20)$$

White *et al.* (1) also assumed that newborns receive a proportion $P_{mat}$ of the anti-parasite and clinical immunity from their mother and that this immunity decays with rate $1/d_{mat}$. Maternally-acquired immunity as a function of time *t*, age *a*, and exposure $\zeta$ is thus modeled as

$$A_{P,mat}(t,a,\zeta)=P_{mat}A_P^*(t-a,20,\zeta)e^{-\frac{a}{d_{mat}}}, \quad (S21)$$

$$A_{C,mat}(t,a,\zeta)=P_{mat}A_C^*(t-a,20,\zeta)e^{-\frac{a}{d_{mat}}}. \quad (S22)$$

In eqs. (S21-S22), $A_P^*(t-a,20,\zeta)$ and $A_C^*(t-a,20,\zeta)$ are the respective anti-parasite and clinical immunity levels of a 20-year-old woman. These quantities are computed as the weighted average across all possible quantities of hypnozoite batches:

$$A_P^*(t-a,20,\zeta)=\sum_{k=1}^{K} Z_H^k(t-a,20,\zeta)A_P^k(t-a,20,\zeta), \quad (S23)$$

$$A_C^*(t-a,20,\zeta) = \sum_{k=1}^{K} Z_H^k(t-a,20,\zeta) A_C^k(t-a,20,\zeta), \qquad (S24)$$

*Effect of Immunity*

White *et al.* (1) then related the levels of anti-parasite and clinical immunity to $\phi_{LM}$, $\phi_D$, and $d_{PCR}$ (i.e., $1/r_{PCR}$) using Hill functions:

$$\phi_{LM}^k(t,a,\zeta) = \phi_{LM,min} + (\phi_{LM,max} - \phi_{LM,min}) \frac{1}{1 + \left(\frac{A_P^k(t,a,\zeta) + A_{P,mat}(t,a,\zeta)}{A_{LM,50\%}}\right)^{\kappa_{LM}}}, \qquad (S25)$$

$$\phi_D^k(t,a,\zeta) = \phi_{D,min} + (\phi_{D,max} - \phi_{D,min}) \frac{1}{1 + \left(\frac{A_C^k(t,a,\zeta) + A_{C,mat}(t,a,\zeta)}{A_{D,50\%}}\right)^{\kappa_C}}, \qquad (S26)$$

$$d_{PCR}^k(t,a,\zeta) = d_{PCR,min} + (d_{PCR,max} - d_{PCR,min}) \frac{1}{1 + \left(\frac{A_P^k(t,a,\zeta) + A_{P,mat}(t,a,\zeta)}{A_{PCR,50\%}}\right)^{\kappa_{PCR}}}, \qquad (S27)$$

*Compartmental Model of P. vivax Transmission in Humans*

Using eqs. (S1-S27), White *et al.* (1) generated the following compartmental model of *P. vivax* transmission in humans:

$$\frac{\partial S^k}{\partial t}+\frac{\partial S^k}{\partial a}+\frac{\partial S^k}{\partial \zeta}$$

$$= -\lambda_H^0(t-d_E,a-d_E,\zeta)S^k - fkS^k + r_{PCR}^k I_{PCR}^k + r_P P^k - \gamma_L k S^k$$

$$+ \gamma_L(k+1)S^{k+1} - \mu_H S^k + \mu_H W(\zeta) I(a=0), \quad (S28)$$

$$\frac{\partial I_{PCR}^k}{\partial t}+\frac{\partial I_{PCR}^k}{\partial a}+\frac{\partial I_{PCR}^k}{\partial \zeta}$$

$$= -\lambda_H^0(t-d_E,a-d_E,\zeta)I_{PCR}^k - fkI_{PCR}^k - r_{PCR}^k I_{PCR}^k + r_{LM} I_{LM}^k$$

$$+ \lambda_H^0(t-d_E,a-d_E,\zeta)\left(1-\phi_{LM}^{k-1}\right)\left(S^{k-1}+I_{PCR}^{k-1}\right) + fk\left(1-\phi_{LM}^k\right)\left(S^k+I_{PCR}^k\right)$$

$$- \gamma_L k I_{PCR}^k + \gamma_L(k+1)I_{PCR}^{k+1} - \mu_H I_{PCR}^k, \quad (S29)$$

$$\frac{\partial I_{LM}^k}{\partial t}+\frac{\partial I_{LM}^k}{\partial a}+\frac{\partial I_{LM}^k}{\partial \zeta}$$

$$= -\lambda_H^0(t-d_E,a-d_E,\zeta)I_{LM}^k - fkI_{LM}^k - r_{LM} I_{LM}^k + r_D I_D^k$$

$$+ \lambda_H^0(t-d_E,a-d_E,\zeta)\left(1-\phi_D^{k-1}\right)\left(\phi_{LM}^{k-1}S^{k-1}+\phi_{LM}^{k-1}I_{PCR}^{k-1}+I_{LM}^{k-1}\right)$$

$$+ fk\left(1-\phi_D^k\right)\left(\phi_{LM}^k S^k+\phi_{LM}^k I_{PCR}^k+I_{LM}^k\right) - \gamma_L k I_{LM}^k + \gamma_L(k+1)I_{LM}^{k+1}$$

$$- \mu_H I_{LM}^k, \quad (S30)$$

$$\frac{\partial I_D^k}{\partial t}+\frac{\partial I_D^k}{\partial a}+\frac{\partial I_D^k}{\partial \zeta}$$

$$= -\lambda_H^0(t-d_E,a-d_E,\zeta)I_D^k + \lambda_H^0(t-d_E,a-d_E,\zeta)I_D^{k-1} - r_D I_D^k$$

$$+ \lambda_H^0(t-d_E,a-d_E,\zeta)\phi_D^{k-1}(1-\chi_T)\left(\phi_{LM}^{k-1}S^{k-1}+\phi_{LM}^{k-1}I_{PCR}^{k-1}+I_{LM}^{k-1}\right)$$

$$+ fk\phi_D^k(1-\chi_T)\left(\phi_{LM}^k S^k+\phi_{LM}^k I_{PCR}^k+I_{LM}^k\right) - \gamma_L k I_D^k + \gamma_L(k+1)I_D^{k+1}$$

$$- \mu_H I_D^k, \quad (S31)$$

$$\frac{\partial T^k}{\partial t}+\frac{\partial T^k}{\partial a}+\frac{\partial T^k}{\partial \zeta}$$

$$= -\lambda_H^0(t-d_E,a-d_E,\zeta)T^k+\lambda_H^0(t-d_E,a-d_E,\zeta)T^{k-1}-r_T T^k$$

$$+\lambda_H^0(t-d_E,a-d_E,\zeta)\phi_D^{k-1}\chi_T\left(\phi_{LM}^{k-1}S^{k-1}+\phi_{LM}^{k-1}I_{PCR}^{k-1}+I_{LM}^{k-1}\right)$$

$$+fk\phi_D^k\chi_T\left(\phi_{LM}^k S^k+\phi_{LM}^k I_{PCR}^k+I_{LM}^k\right)-\gamma_L kT^k+\gamma_L(k+1)T^{k+1}$$

$$-\mu_H T^k, \quad (S31)$$

$$\frac{\partial P^k}{\partial t}+\frac{\partial P^k}{\partial a}+\frac{\partial P^k}{\partial \zeta}$$

$$= -\lambda_H^0(t-d_E,a-d_E,\zeta)P^k+\lambda_H^0(t-d_E,a-d_E,\zeta)P^{k-1}+r_T T^k-r_P P^k$$

$$-\gamma_L kP^k+\gamma_L(k+1)P^{k+1}-\mu_H P^k, \quad (S32)$$

*Compartmental Model of P. vivax Transmission in Mosquitoes*

White *et al.* (1) also explicitly modeled the population dynamics of the *Anopheles* vector and the transmission dynamics therein. For a given *Anopheles* species *v*, the force of infection experienced by the mosquito population is calculated as

$$\lambda_M^v(t)=\alpha^v\int_\zeta\int_a \zeta X(a)(c_{PCR}I_{PCR}(t,a,\zeta)+c_{LM}I_{LM}(t,a,\zeta)+c_D I_D(t,a,\zeta)$$

$$+c_T I_T(t,a,\zeta))\,da\,d\zeta. \quad (S33)$$

In eq. (S33), $c_{PCR}$, $c_{LM}$, $c_D$, and $c_T$ represent the probabilities of infection to mosquitoes from humans during PCR-detectable, LM-detectable, clinical, or clinical and treated infection, respectively.

The compartmental model tracks the mosquito population from early instar larvae ($L_E^v$) to late instar larvae ($L_L^v$) and pupae ($L_P^v$) as well as susceptible ($S_M^v$), exposed ($E_M^v$), and infected ($I_M^v$) adult mosquitoes. The system of differential equations regulating the transition through these compartments is

$$\frac{dL_E^v}{dt} = \beta^v m^v - \mu_E^{0,v}\left(1 + \frac{L_E^v + L_L^v}{K^v(t)}\right)L_E^v - \frac{L_E^v}{d_E^v}, \quad (S34)$$

$$\frac{dL_L^v}{dt} = \frac{L_E^v}{d_E^v} - \mu_L^{0,v}\left(1 + \gamma^v\frac{L_E^v + L_L^v}{K^v(t)}\right)L_L^v - \frac{L_L^v}{d_L^v}, \quad (S35)$$

$$\frac{dL_P^v}{dt} = \frac{L_P^v}{d_L^v} - \mu_P^v L_P^v - \frac{L_P^v}{d_P^v}, \quad (S36)$$

$$\frac{dS_M^v}{dt} = \frac{1}{2}\frac{L_P^v}{d_P^v} - \lambda_M^v(t)S_M^v - \mu_M^v S_M^v, \quad (S37)$$

$$\frac{dE_M^v}{dt} = \lambda_M^v(t)S_M^v - \lambda_M^v(t-\tau_M^v)e^{-\mu_M^v\tau_M^v}S_M^v(t-\tau_M^v) - \mu_M^v E_M^v, \quad (S38)$$

$$\frac{dI_M^v}{dt} = \lambda_M^v(t-\tau_M^v)e^{-\mu_M^v\tau_M^v}S_M^v(t-\tau_M^v) - \mu_M^v I_M^v, \quad (S39)$$

The parameters and their assumed values governing the system of differential equations can be found in Table S3.

***Table S1. Default Transmission Setting Parameters for the Human Component of the Transmission Model.*** *A description of the default parameters used in the individual-based model to simulate clinical trial data are provided along with the numeric values and the corresponding references.*

| Description | Parameter | Value | Reference |
|---|---|---|---|
| **Demographics** | | | |
| Mean Age of Population | $1/\mu_H$ | 31.8 years | Assumed |
| Max Age of Population | $a_{max}$ | 80 years | Assumed |
| Proportion of Pregnant Women | | 7.5% | Assumed |
| G6PD Deficiency Prevalence | $q_{G6PD}$ | 0% | Assumed |
| CYP-2D6 Phenotype Prevalence | $q_{CYP2D6}$ | 0% | Assumed |
| **Exposure to Mosquito Biting** | | | |
| Degree of Age-Dependent Biting | $\rho_{age}$ | 0.85 | (3,4) |
| Reference Age of Biting | $a_0$ | 8 years | (3,4) |
| Mosquito-to-Human Transmission Probability | $b$ | 0.50 | (2) |
| **Hypnozoite Dynamics** | | | |
| Rate of Relapse | $f$ | 1/65 $day^{-1}$ | (2) |
| Rate of Liver-Stage Hypnozoite Clearance | $\gamma_L$ | 1/383 $day^{-1}$ | (1) |

***Table S2. Default Infection Dynamics Parameters for the Human Component of the Transmission Model.*** *A description of the default parameters used in the individual-based model to simulate clinical trial data are provided along with the numeric values and the corresponding references.*

| Description | Parameter | Value | Reference |
|---|---|---|---|
| **Human Infection Duration** | | | |
| Latent Period | $d_E$ | 10 days | (5) |
| Max PCR-Detectable Infection (No Immunity) | $d_{PCR,max}$ | 70 days | (1) |
| Min PCR-Detectable Infection (Full Immunity) | $d_{PCR,min}$ | 10 days | (1) |
| LM-Detectable Infection | $d_{LM}$ | 10 days | (2) |
| Clinical Disease (Untreated) | $1/r_D$ | 5 days | (1) |
| Clinical Disease (Treated) | $1/r_T$ | 1 day | (1) |
| **Immunity** | | | |
| Anti-Parasite Immune Boosting Refractory Period | $u_{par}$ | 19.77 days | (2) |
| Duration of Anti-Parasite Immunity | $d_{par}$ | 10 years | (1) |
| Probability of LM-Detectable Infection (No Immunity) | $\phi_{LM,max}$ | 0.8918 | (2) |
| Probability of LM-Detectable Infection (Full Immunity) | $\phi_{LM,min}$ | 0.0043 | (2) |
| Anti-Parasite Immunity for 50% Reduction in Duration of LM-Detectable Infection | $A_{LM,50\%}$ | 27.52 | (2) |
| Shape Parameter for LM-Detectable Infection | $\kappa_{LM}$ | 2.403 | (2) |
| Anti-Parasite Immunity for 50% Reduction in Duration of PCR-Detectable Infection | $A_{PCR,50\%}$ | 9.9 | (2) |
| Shape Parameter for Duration of PCR-Detectable Infection | $\kappa_{PCR}$ | 4.602 | (2) |
| Clinical Immune Boosting Refractory Period | $u_{clin}$ | 7.85 days | (2) |
| Duration of Clinical Immunity | $d_{clin}$ | 30 years | (1) |
| Probability of Clinical Episode (No Immunity) | $\phi_{D,max}$ | 0.8605 | (2) |
| Probability of Clinical Episode (Full Immunity) | $\phi_{D,min}$ | 0.018 | (2) |
| Clinical Immunity for 50% Reduction in Clinical Episode | $A_{D,50\%}$ | 11.538 | (2) |
| Shape Parameter for Probability of Clinical Episode | $\kappa_D$ | 2.25 | (2) |
| Newborn Immunity Relative to Mother's | $P_{mat}$ | 0.421 | (2) |
| Duration of Maternal Immunity | $d_{mat}$ | 35.148 | (2) |
| **Infectiousness to Mosquitoes** | | | |
| During PCR-Detectable Infection | $c_{PCR}$ | 0.035 | (6) |
| During LM-Detectable Infection | $c_{LM}$ | 0.1 | (6) |
| During Clinical Disease | $c_D$ | 0.8 | (6) |
| During Treated Clinical Disease | $c_T$ | 0.4 | (6) |

***Table S3. Default Parameters for the Mosquito Component of the Transmission Model.*** *A description of the default parameters used in the individual-based model to simulate clinical trial data are provided along with the numeric values and the corresponding reference.*

| Description | Parameter | Value | Reference |
|---|---|---|---|
| **Mosquito Bionomics** | | | |
| Mosquito Life Expectancy | $1/\mu_M^v$ | 6 days | (2) |
| Duration of Sporogony | $\tau_M^v$ | 8.4 days | (2) |
| Human Blood Index | $Q_0^v$ | 0.50 | (2) |
| Proportion of Bites Occurring in Bed | $\Phi_B$ | 0.45 | Assumed |
| Proportion of Bites Occurring Indoors | $\Phi_I$ | 0.90 | Assumed |
| **Mosquito Larval Development** | | | |
| Development Time of Early Larval Instars | $d_E^v$ | 6.64 days | (7) |
| Development Time of Late Larval Instars | $d_L^v$ | 3.72 days | (7) |
| Development Time of Pupae | $d_P^v$ | 0.64 days | (7) |
| Per Capita Mortality Rate of Early Larval Instars | $\mu_E^{0,v}$ | 0.034 day$^{-1}$ | (7) |
| Per Capita Mortality Rate of Late Larval Instars | $\mu_L^{0,v}$ | 0.035 day$^{-1}$ | (7) |
| Per Capita Mortality Rate of Pupae | $\mu_P^v$ | 0.25 day$^{-1}$ | (7) |
| Daily Number of Eggs Laid per Mosquito | $\beta^v$ | 21.19 | (7) |
| Effect of Density Dependence on Late Larval Instars Relative to Early Larval Instars | $\gamma^v$ | 13.25 | (7) |

***Trial Design***

*Power Calculations*

To ensure that our simulated clinical trials were appropriately powered, we performed a retrospective power analysis. We took the outputs of our simulated clinical trials performed in transmission settings with homogeneous biting and computed the median and interquartile range of sample sizes in each arm necessary to achieve a specific level of power at a given EIR. The incidence rate and the proportion at risk in the control arm were computed using the full record of LM-detectable/subclinical and clinical relapses caused by hypnozoites acquired prior to treatment. For each power calculation, we assumed a type-I error rate of 0.05 and an efficacy of 75%.

Using the Cox proportional hazards model (8), we first calculated the total number of events $N$ needed to have power $\beta$ and type-I error rate $\alpha$ as

$$N = \frac{\left(z_\beta + z_{1-\frac{\alpha}{2}}\right)^2}{P_t P_c \log^2(E_{Cox})}, \quad (S40)$$

where $z_\beta$ and $z_{1-\frac{\alpha}{2}}$ are the z-scores, $P_t$ and $P_c$ are the respective proportion of participants in the treatment and control arms, and $E_{Cox}$ is the expected efficacy. We then calculated the number of participants in the control arm by dividing the total number of events needed by the expected total incidence rate computed from the trial output. The number of participants in the treatment arm was equal to the number of participants in the control arm.

Based on incidence rates, we followed Smith *et al.* (9) and computed the number of participants in each arm as

$$n = \left(z_\beta + z_{1-\frac{\alpha}{2}}\right)^2 \frac{r_c + r_t}{(r_c - r_t)^2}, \quad (S41)$$

where $z_\beta$ and $z_{1-\frac{\alpha}{2}}$ are the z-scores, $r_c$ is the expected incidence rate in the control arm, and $r_t$ is the expected incidence rate in the treatment arm.

Based on proportion at risk, we computed the number of participants in each arm as

$$n = \left(z_\beta + z_{1-\frac{\alpha}{2}}\right)^2 \frac{2p(1-p)}{(p_c - p_t)^2}. \quad (S42)$$

In eq. (S42), $z_\beta$ and $z_{1-\frac{\alpha}{2}}$ are the z-scores, $p_t$ and $p_c$ are the expected proportion of participants at risk in the treatment and control arms, and $p$ is the average of $p_t$ and $p_c$ (9).

### ***Simulation Scenarios***

*Effect of Transmission Intensity and Heterogeneity in Biting*

The transmission intensity at the trial location may determine the frequency, cause, and detectability of recurrent infections that occur within the duration of follow-up. The number of reinfection events experienced by each trial participant increases with EIR. Thus, at higher EIR, the proportion of recurrent infections that are relapses caused by hypnozoites acquired prior to treatment may be small. Moreover, the levels of anti-parasite and clinical immunity in each trial participant is a function of exposure and transmission intensity, and, as EIR increases, fewer recurrent infections may be detected by light microscopy. Therefore, the transmission intensity at the trial location may bias efficacy estimates, likely downward, due to the rate at which trial participants are reinfected by mosquitoes.

Heterogeneity in mosquito biting contributes to observed heterogeneous patterns of *Plasmodium* transmission (10). For *P. falciparum*, heterogeneity in biting biases efficacy estimates downward, because fewer participants in the treatment and control arms are exposed within the duration of follow-up (11). For *P. vivax*, however, heterogeneity in biting may reduce the bias due to transmission intensity, because fewer participants are reinfected by mosquitoes within the duration of follow-up.

We varied the transmission intensity and the degree of heterogeneity in mosquito biting to quantify their effect on efficacy estimates. We varied transmission intensity by simulating an equilibrium population-level EIR of 1, 10, or 100 infectious bites per person-year (*ibppy*), consistent with the range of values reported by White *et al.* (1). To account for heterogeneity in biting, we varied the variance of the distribution of logged biting propensities from zero to three. This distribution determines the variation in exposure to mosquito biting across individuals, and a zero variance implies homogeneous biting.

*Effect of Rate of Relapse and Duration of Follow-up*

The rate of *P. vivax* relapse varies geographically, with the estimated mean time to first relapse ranging from 41-65 days for tropical phenotypes to 299 days for temperate phenotypes (12). If the duration of follow-up is too short relative to the mean time to first relapse, then an appreciable number of trial participants may not have relapsed by the duration of follow-up. A longer duration of follow-up may capture a great proportion of relapses but also increases the temporal window over which trial participants can become reinfected by mosquito biting. The probability of reinfection during the duration of follow-up will vary with the EIR.

We examined the interaction between the rate of relapse, the duration of follow-up, and transmission intensity and quantified its effect on the efficacy estimates in simulated clinical trials. We fixed the mean time to relapse at 30, 60, 90, or 180 days and simulated clinical trials in which the duration of follow-up was 90, 180, 365, or 730 days. The mean times to relapse chosen are consistent with the range of estimates reported in Battle *et al*. (12). Trial participants were assessed for asexual parasites using light microscopy according to the following schedule: days 8, 15, 22, 29, 60, and 90 for the 90-day follow-up period; days 8, 15, 22, 29, 60, 90, 120, and 180 for the 180-day follow-up period; days 8, 15, 22, 29, 60, 90, 120, 180, 240, 300, and 365 for the 365-day follow-up period; and days 8, 15, 22, 29, 60, 90, 120, 180, 240, 300, 360, 420, 480, 540, 600, 660, and 730 for the 730-day follow-up period. Moreover, to examine the effect of transmission intensity, we set the population-level EIR at 1, 10, or 100. To isolate the joint effect of the rate of relapse and duration of follow-up across transmission intensities, we assumed homogeneous biting.

*Effect of Vector Control*

The implementation of vector control measures may protect trial participants against mosquito biting and reduce the number of reinfection events during follow-up. As a result, vector control may reduce the bias due to frequent reinfection of trial participants at higher transmission intensities.

To evaluate whether vector control measures are effective in reducing the bias in the efficacy estimates, we simulated clinical trials under four different vector control scenarios: (1) no vector control; (2) distribution of LLINs; (3) administration of IRS; and (4) distribution of LLINs and administration of IRS. The extent to which LLINs and IRS prevent mosquito biting depends upon the biting behavior of the mosquito population. We simulated different biting behaviors by varying the absolute proportion of bites that occurred while individuals were indoors ($\Phi_I$) and while individuals were in bed ($\Phi_B$). Greater values of $\Phi_I$ correspond to greater endophagy (i.e., indoor feeding), and greater values of ${}^{\Phi_B}/_{\Phi_I}$ correspond to a greater preference towards nighttime biting. We considered four biting behaviors: (1) endophagic with no preference in biting time ($\Phi_I = 0.9;\ \Phi_B = 0.45$); (2) endophagic with nighttime biting ($\Phi_I = 0.9;\ \Phi_B = 0.675$); (3) endophagic with daytime biting ($\Phi_I = 0.9; \Phi_B = 0.225$); and (4) exophagic with no preference in biting time ($\Phi_I = 0.1; \Phi_B = 0.05$). We simulated each combination of vector control scenario and mosquito biting behavior at population-level EIRs of 1, 10, and 100. By default, we assumed homogeneous biting.

*Effect of Genotyping*

Although vector control may decrease the bias by reducing the number of reinfection events, a method capable of distinguishing relapses caused by hypnozoites acquired prior to treatment from all other recurrent infections is needed to completely correct the bias in efficacy estimates. Methods leveraging time-to-event and genotyping data have been successfully used to determine the cause of each *P. vivax* recurrent infection (13,14) and therefore could reduce and, under certain circumstances, possibly correct biases in phase-III clinical trials.

We evaluated the potential for such a method at varying sensitivities and specificities to correct the bias in the efficacy estimates. We considered a method intended to distinguish relapses caused from hypnozoite batches acquired prior to treatment from all other recurrent infections. Therefore, the sensitivity of the method was the probability of correctly identifying a relapse caused by a hypnozoite batch acquired prior to treatment, and the specificity of the method was the probability of correctly identifying all other recurrent infections. We simulated clinical trials at equilibrium population-level EIRs of 1, 10, and 100, assuming homogeneous biting. We then analyzed the simulated outputs of the clinical trials using a hypothetical method with sensitivities and specificities of 25%, 50%, 75%, or 100%.

*Effect of the 8-aminoquinoline*

By default, we simulated clinical trials for an 8-aminoquinoline with prophylactic effect lasting 28 days, consistent with treatment with chloroquine and primaquine. Under this assumption, the duration of prophylaxis is less than the 32-day period of left-censoring, so the suppression of blood-stage parasites during prophylaxis will not bias our efficacy estimates. Treatment with tafenoquine when co-administered with chloroquine, however, provides prophylaxis for 45 days and, therefore, may bias upwards our efficacy estimates (15).

To examine whether the duration of prophylaxis affects our estimates of efficacy, we simulated and compared phase-III clinical trials for primaquine and tafenoquine with prophylactic effects lasting 28 and 45 days, respectively. Because the benefit of prophylaxis may vary with transmission intensity, we simulated each trial at population-level EIRs of 1, 10, and 100 with homogeneous biting.

*Effect of the efficacy metric and infection endpoint*

The combination of efficacy metric and infection endpoint used to calculate efficacy varied across recent clinical trials (15–18). Different combinations of efficacy metrics and infection endpoints could make comparison of efficacy estimates across different trials challenging if efficacy estimates are sensitive to this feature of the trial design.

To test whether our efficacy estimates are sensitive to the choice of the efficacy metric and the infection endpoint used, we simulated phase-III clinical trials at EIRs of 1, 10, and 100 and calculated efficacy using different combinations of efficacy metrics and infection endpoints. The efficacy metrics examined were the Cox proportional hazards model, incidence rates, and the proportion at risk. The infection endpoints examined were PCR-detectable recurrent infections, LM-detectable recurrent infections, and clinical recurrent infections. Because we wanted to isolate the effect of the efficacy metric and the infection endpoint, we simulated each trial assuming homogeneous biting.

### Results

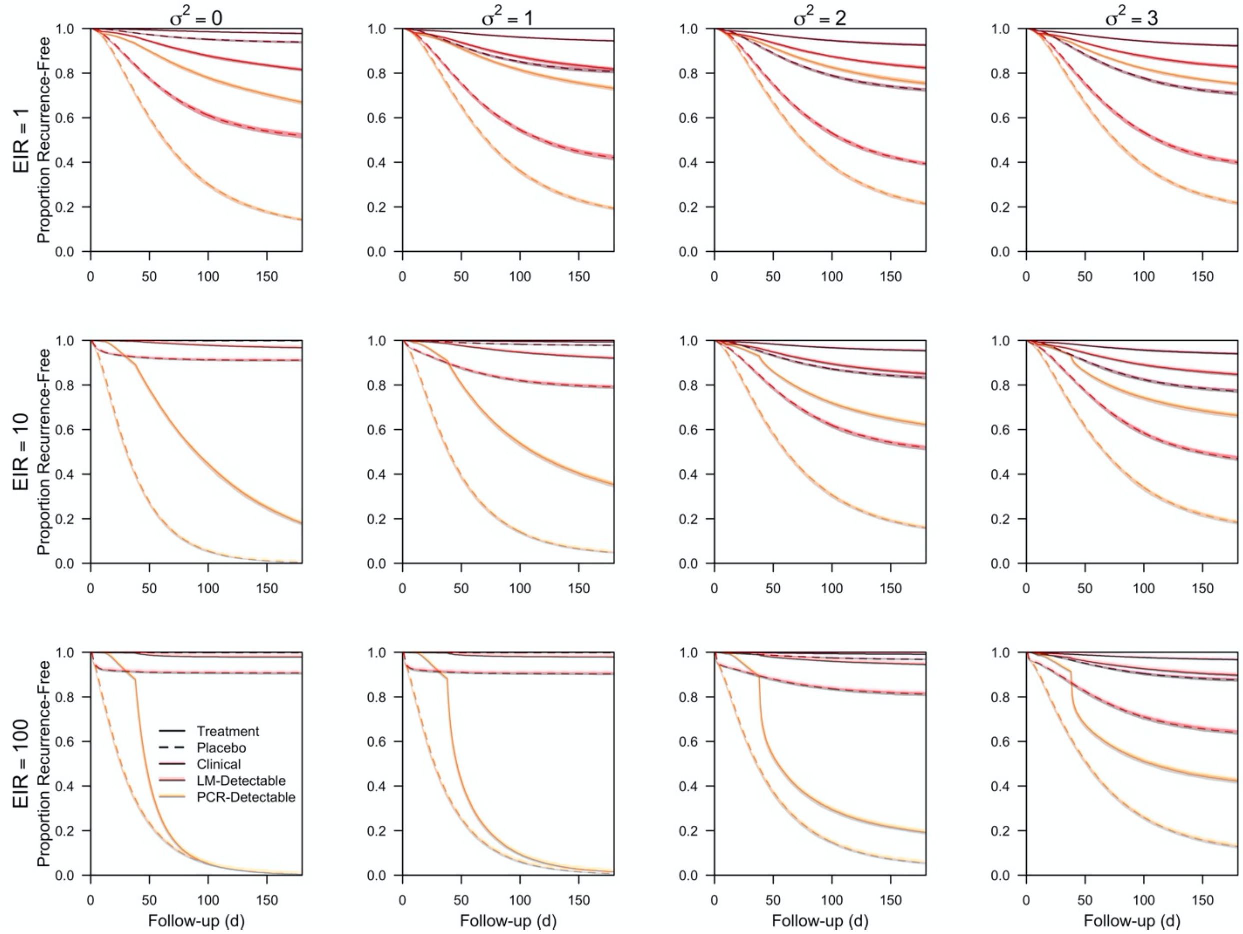


***Figure S1. Recurrence-free survival curves for the effect of transmission intensity and heterogeneity in biting.*** *Recurrence-free survival from simulated clinical trials as a function of follow-up time is shown at different entomological inoculation rates (EIR) and levels of heterogeneity in biting ($\sigma^2$). Each line is in the median of 200 simulations, and each shaded region is the interquartile range. Solid lines correspond to the treatment arm, and dashed lines correspond to the placebo arm. The color of each survival curve corresponds to the infection endpoint used, with orange corresponding to all PCR-detectable recurrent infections, red corresponding to all LM-detectable recurrent infections, and maroon corresponding to all clinical recurrent infections.*

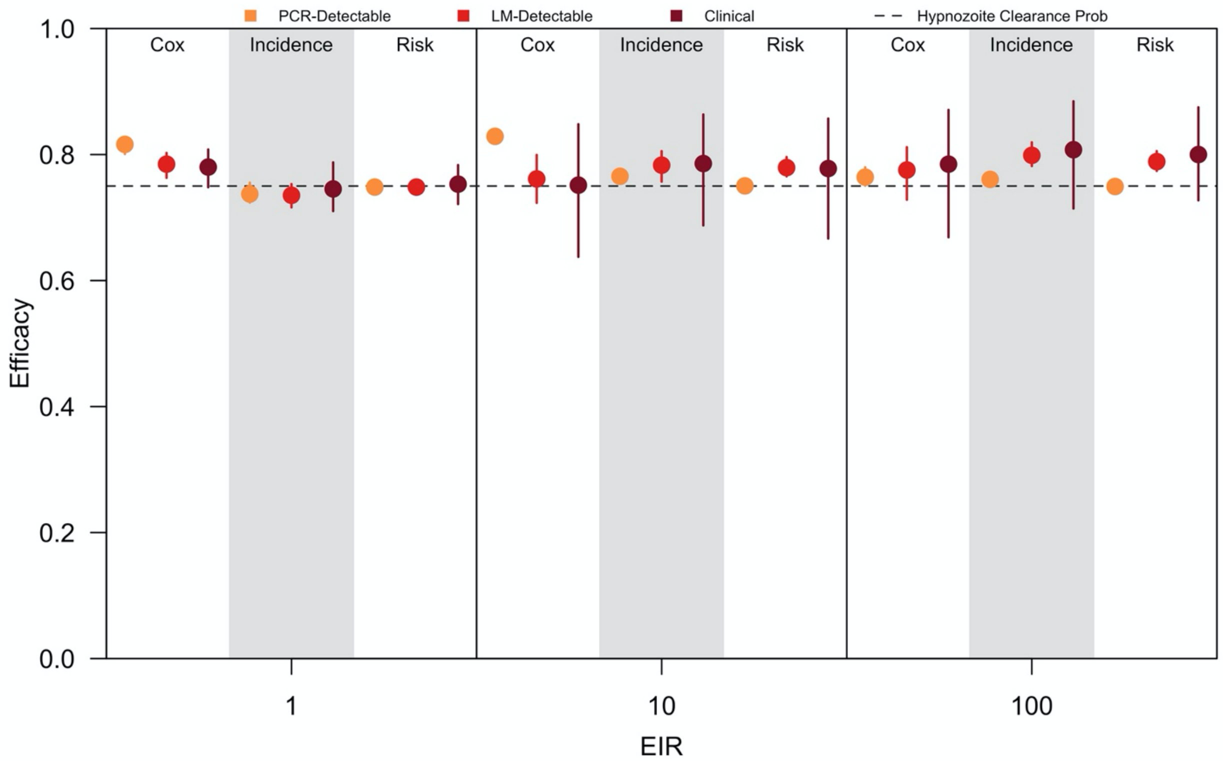


***Fig S2. Effect of efficacy metric and infection endpoint on efficacy estimates under perfect genotyping and complete observation of infections.*** *Efficacy estimates obtained from simulated clinical trials at different entomological inoculation rates (EIR) is shown when calculated using the Cox proportional hazards model, incidence rates, or the proportion at risk, assuming perfect genotyping and complete observation of cases. The infection endpoint was clinical (maroon), LM-detectable (red), or PCR-detectable (orange) relapses caused by hypnozoite batches acquired prior to treatment and identified during follow-up. Each point is the median of 200 simulations, and each bar is the interquartile range. The dotted line is the clearance probability simulated in the trial.*

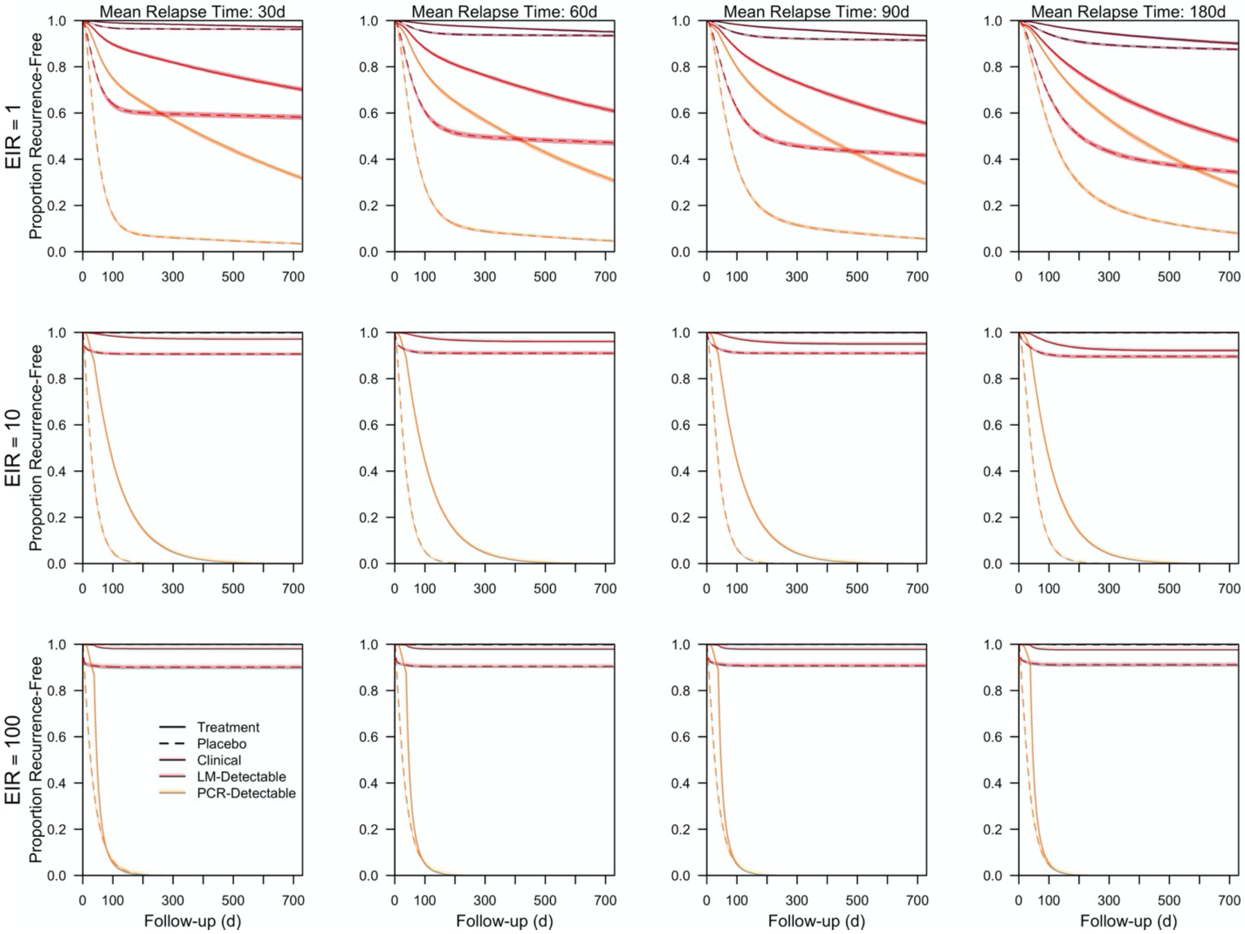


***Figure S3. Recurrence-free survival curves for the effect of rate of relapse and duration of follow-up.*** *Recurrence-free survival curves from simulated clinical trials as a function of follow-up time are show at different entomological inoculation rates (EIR) and mean times to relapse. Each line is the median of 200 simulations, and each shaded region is the interquartile range. Solid lines correspond to the treatment arm, and dashed lines correspond to the placebo arm. The color of each survival curve corresponds to the infection endpoint used, with orange corresponding to all PCR-detectable recurrent infections, red corresponding to all LM-detectable recurrent infections, and maroon corresponding to all clinical recurrent infections.*

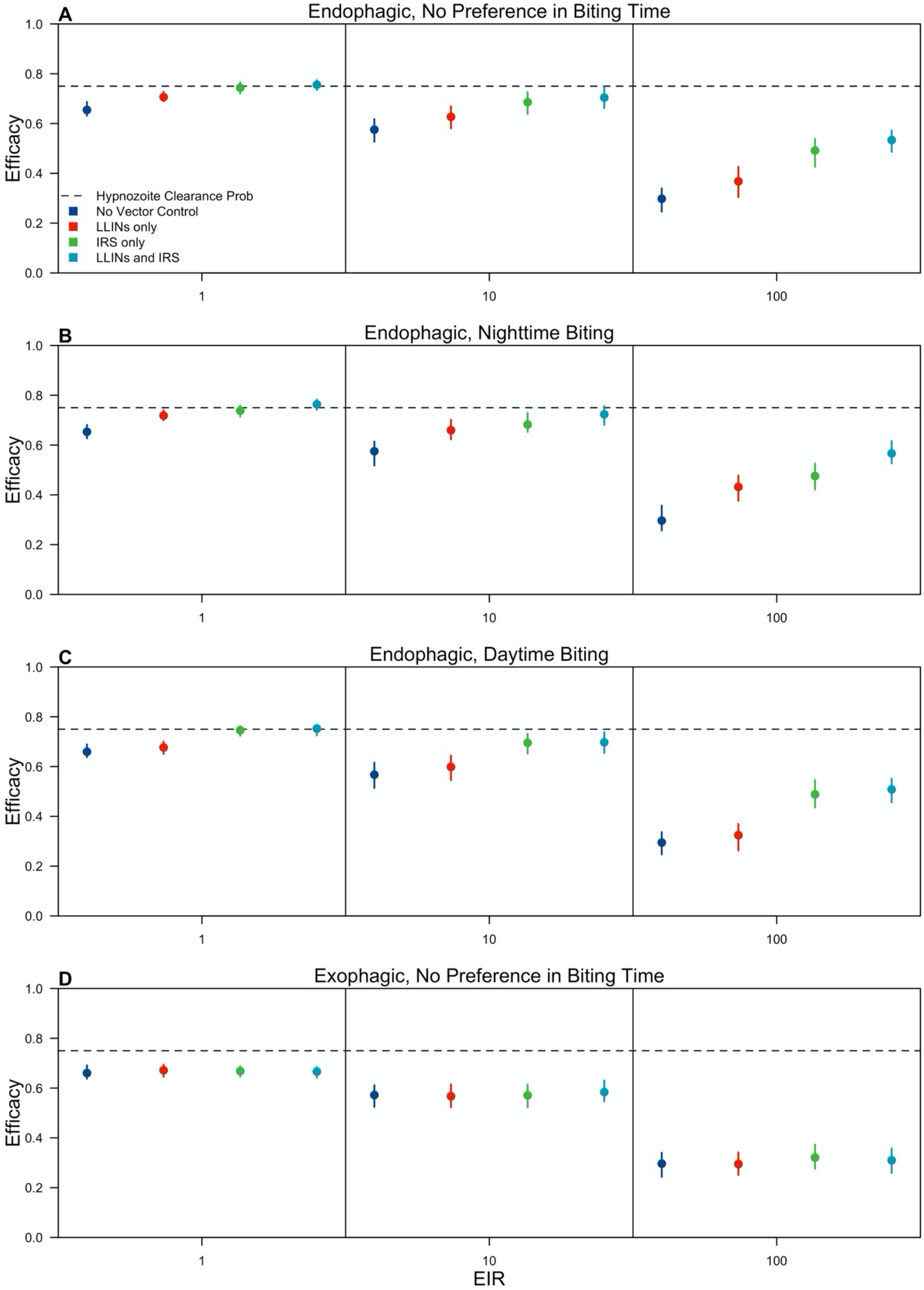
A
Endophagic, No Preference in Biting Time
B
Endophagic, Nighttime Biting
C
Endophagic, Daytime Biting
D
Exophagic, No Preference in Biting Time
Efficacy
EIR
1
10
100
1.0
0.8
0.6
0.4
0.2
0.0
Hypnozoite Clearance Prob
No Vector Control
LLINs only
IRS only
LLINs and IRS

413

***Fig S4. Effect of vector control on efficacy estimates across different vector bionomics.*** *The impact of LLIN distribution (red), IRS administration (green), and combined LLIN distribution and IRS administration (teal) on LM-detectable recurrence free efficacy estimates is compared to a no-intervention scenario (dark blue) across a range of entomological inoculation rates (EIR). Each point represents the median of 200 simulations, and each bar is the interquartile range. The dotted line is the clearance probability simulated in each trial. The absolute proportion of bites occurring indoors* ($\Phi_I$) *and in bed* ($\Phi_B$) *varied with (A)* $\Phi_I = 0.9$ *and* $\Phi_B = 0.45$*; (B)* $\Phi_I = 0.9$ *and* $\Phi_B = 0.675$*; (C)* $\Phi_I = 0.9$ *and* $\Phi_B = 0.225$*; and (D)* $\Phi_I = 0.1$ *and* $\Phi_B = 0.05$

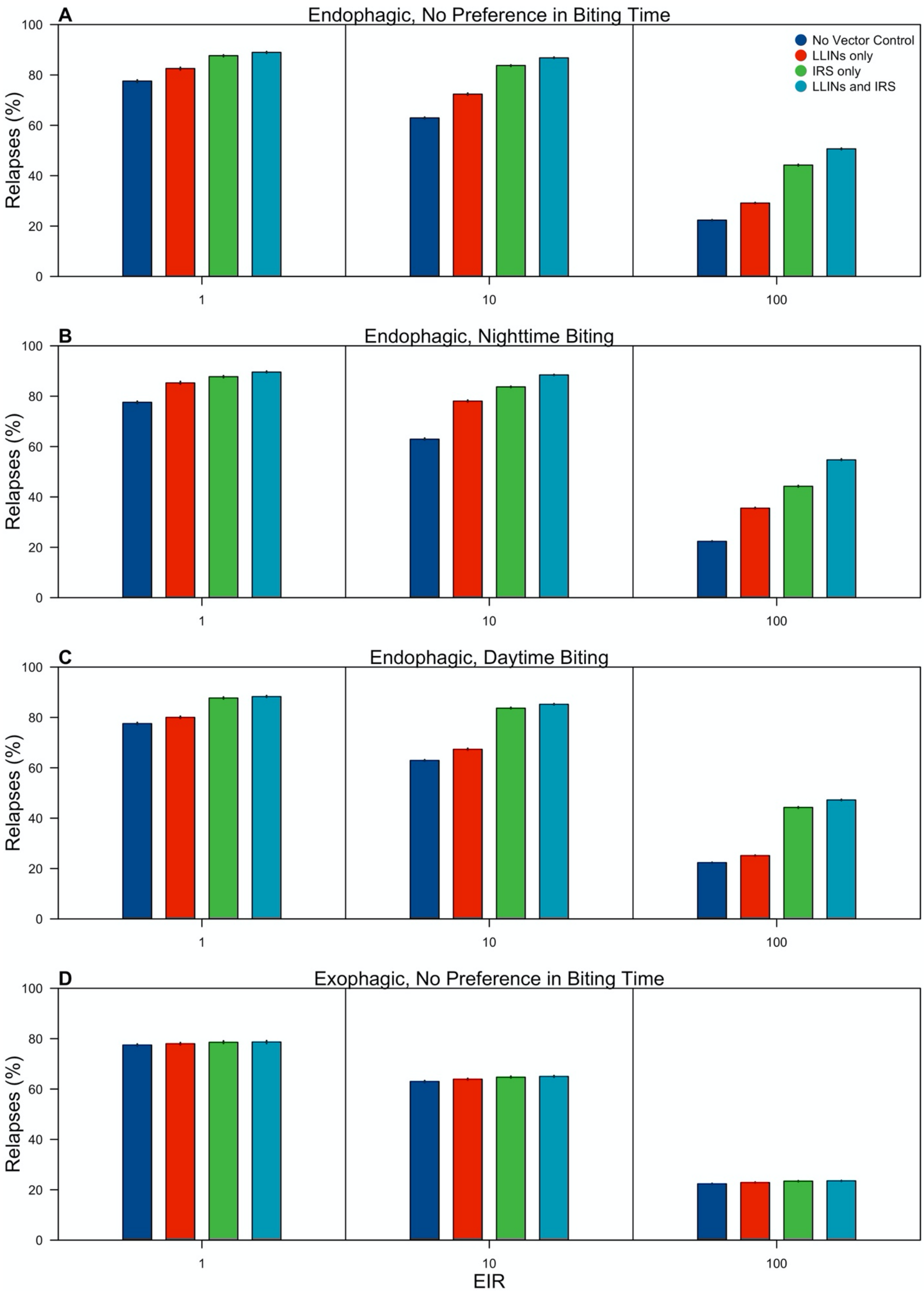

A
Endophagic, No Preference in Biting Time
B
Endophagic, Nighttime Biting
C
Endophagic, Daytime Biting
D
Exophagic, No Preference in Biting Time
Relapses (%)
No Vector Control
LLINs only
IRS only
LLINs and IRS
0
20
40
60
80
100
1
10
100
EIR

***Fig S5. Effect of vector control on the composition of recurrent infections across different vector bionomics.*** *The impact of LLIN distribution (red), IRS administration (green), and combined LLIN distribution and IRS administration (teal) on the percentage of recurrent infections in the control arm that are treatment failures (i.e., relapses caused by hypnozoite batched acquired prior to treatment) is compared to a non-intervention scenario (dark blue) across a range of entomological inoculation rates (EIR). The height of each bar represents the median of 200 simulations, and the segment is the interquartile range. The absolute proportion of bites occurring indoors ($\Phi_I$) and in bed ($\Phi_B$) varied with (A)* $\Phi_I = 0.9$ *and* $\Phi_B = 0.45$*; (B)* $\Phi_I = 0.9$ *and* $\Phi_B = 0.675$*; (C)* $\Phi_I = 0.9$ *and* $\Phi_B = 0.225$*; and (D)* $\Phi_I = 0.1$ *and* $\Phi_B = 0.05$

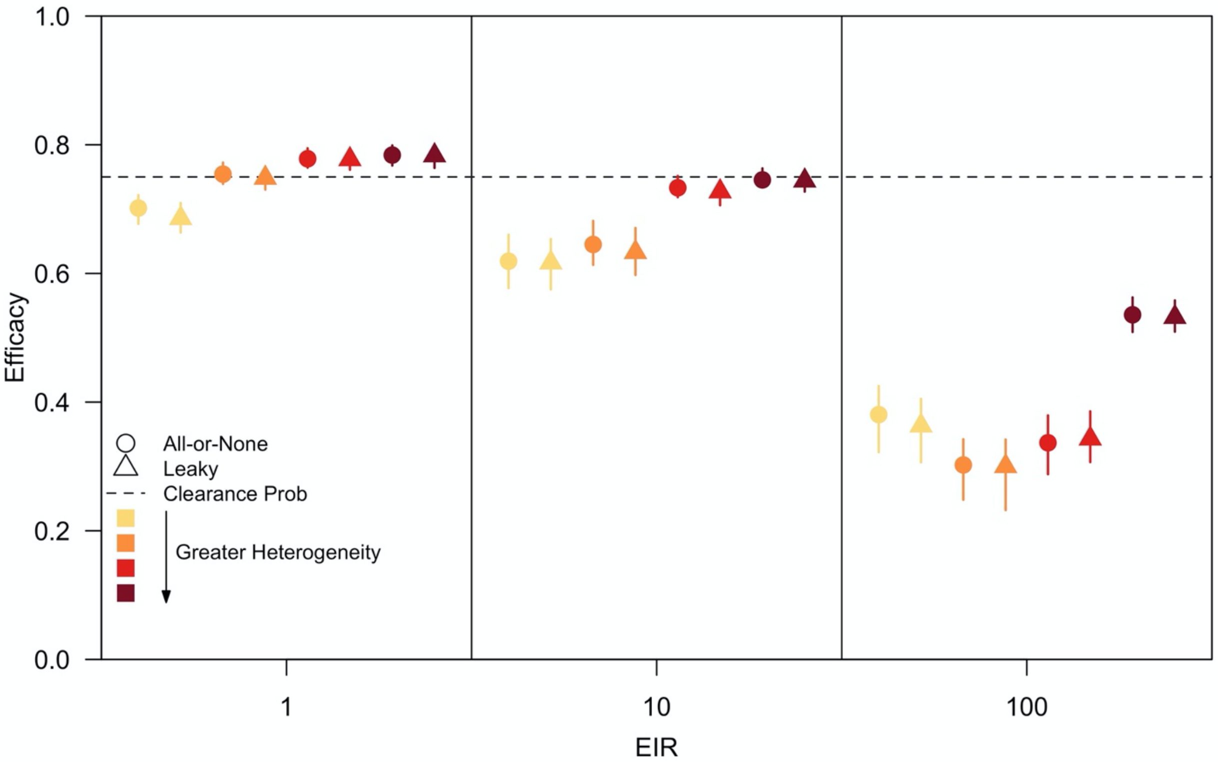


***Fig S6. Effect of transmission intensity and heterogeneity in biting on efficacy estimates for "all-or-none" and "leaky" intervention actions.*** *LM-detectable recurrence-free efficacy estimated from simulated clinical trials is show at different entomological inoculation rates (EIR) and levels of heterogeneity in biting for "all-or-none" (circles) and "leaky" (triangles) intervention actions. Each point represents the median of 200 simulations, and each bar is the interquartile range. The color represents the degree of heterogeneity in individual-level exposure to biting. Darker colors indicate greater heterogeneity in individual-level exposure to biting, and the dotted line is the clearance probability.*

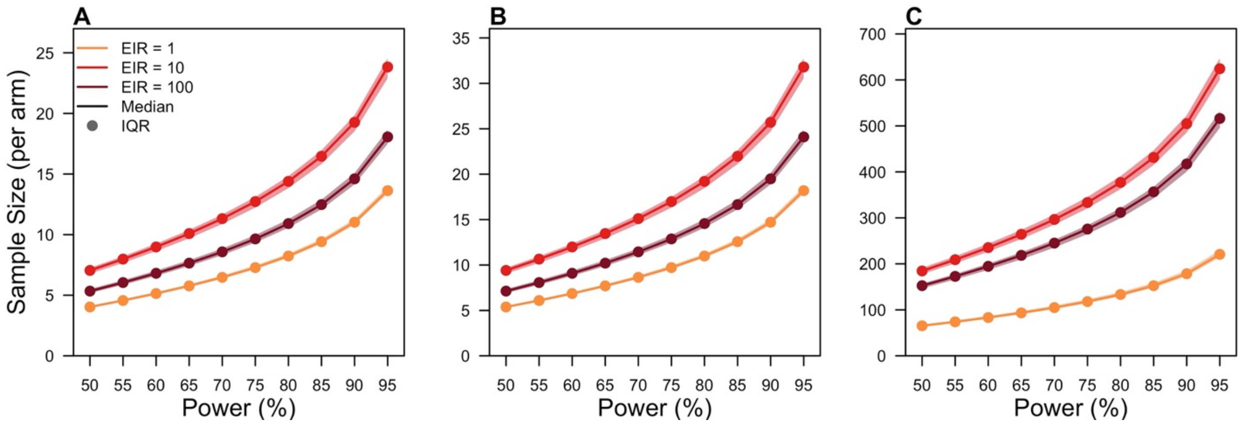


***Fig S7. Sample sizes necessary to achieve a specific level of power.*** *For a given EIR, the sample size in each arm is reported to achieve a specific level of power when calculating efficacy using (A) the Cox proportional hazards model, (B) incidence rates, and (C) the proportion at risk. Incidence rates were computed using the number of relapses caused by hypnozoites acquired prior to treatment in the control arm. Power calculations were performed for each of 200 simulations assuming homogeneous biting. The points and line denote the median sample size, and the shaded region is the interquartile range. Efficacy of the intervention was assumed to be 75%, the type-I error rate was set to 0.05, and the allocation ratio between trial arms was one.*

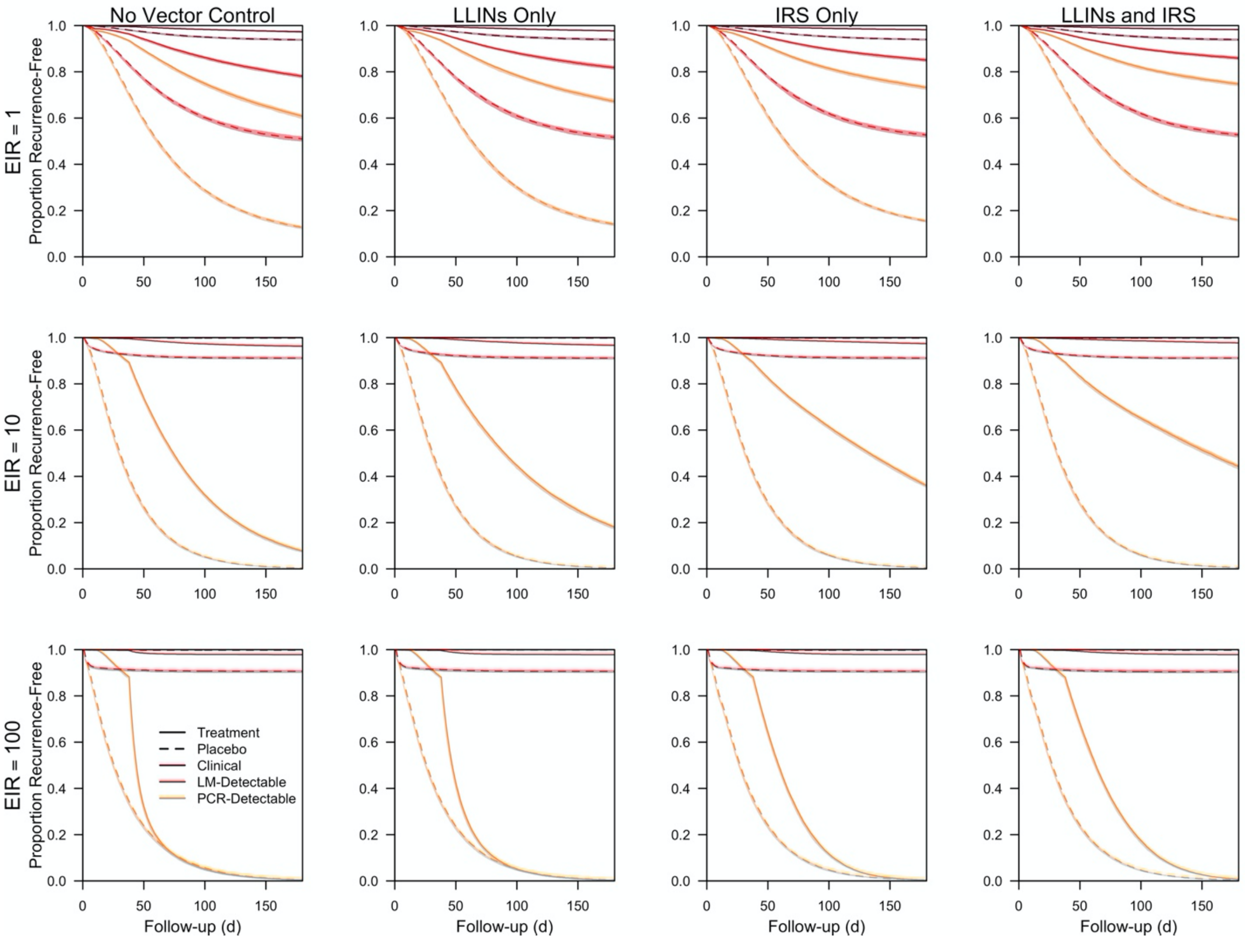

***Figure S8. Recurrence-free survival curves for the effect of vector control.*** *Recurrence-free survival curves from simulated clinical trials as a function of follow-up time are shown at different entomological inoculation rates (EIR) and vector control scenarios. The vector control scenarios considered were no vector control, LLIN distribution, IRS administration, and combined LLIN distribution and IRS administration. Each line is the median of 200 simulations, and each shaded region is the interquartile range. Solid lines correspond to the treatment arm, and dashed lines correspond to the placebo arm. The color of each survival curve corresponds to the infection endpoint used, with orange corresponding to all PCR-detectable recurrent infections, red corresponding to all LM-detectable recurrent infections, and maroon corresponding to all clinical recurrent infections.*

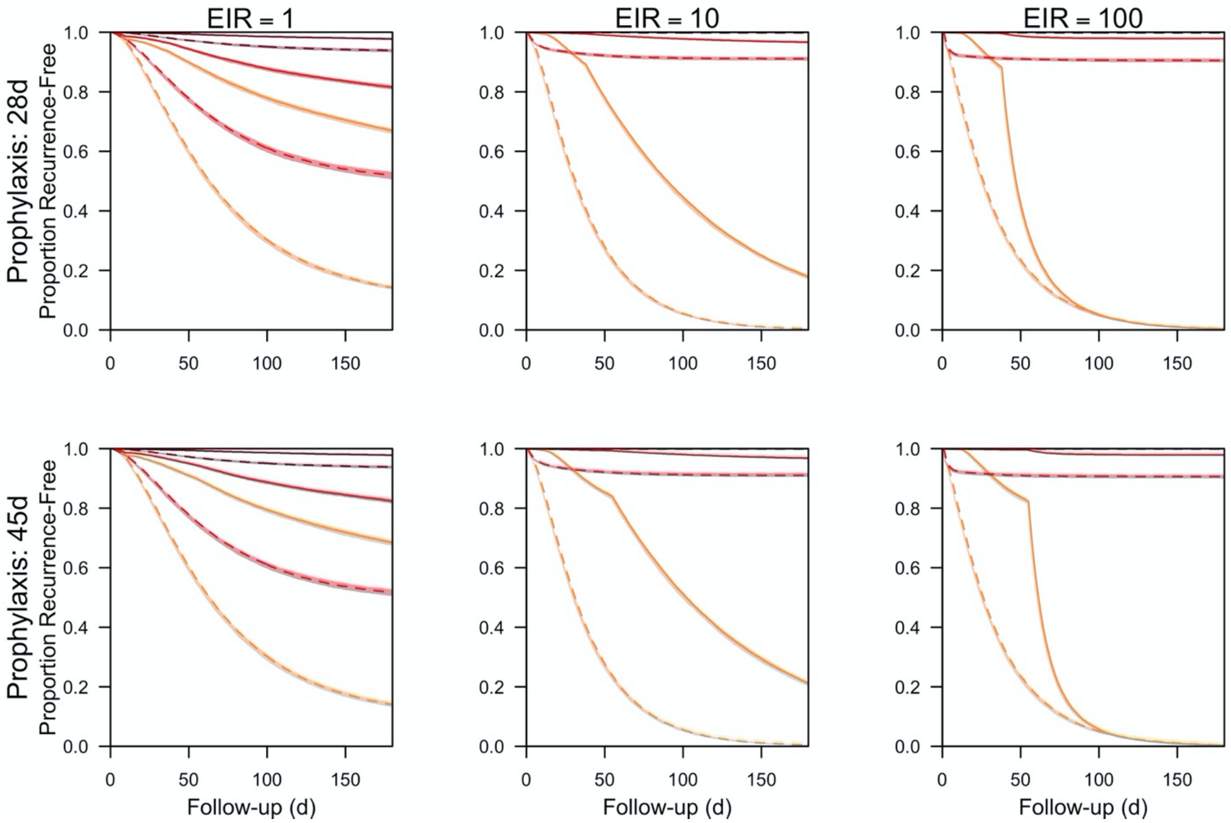


***Figure S9. Recurrence-free survival curves for the effect of 8-aminoquinoline.*** *Recurrence-free survival curves from simulated clinical trials as a function of follow-up time are shown at different entomological inoculation rates (EIR) and 8-aminoquinolines (i.e., primaquine and tafenoquine). Each line is the median of 200 simulations, and each shaded region is the interquartile range. Solid lines correspond to the treatment arm, and dashed lines correspond to the placebo arm. The color of each survival curve corresponds to the infection endpoint used, with orange corresponding to all PCR-detectable recurrent infections, red corresponding to all LM-detectable recurrent infections, and maroon corresponding to all clinical recurrent infections.*

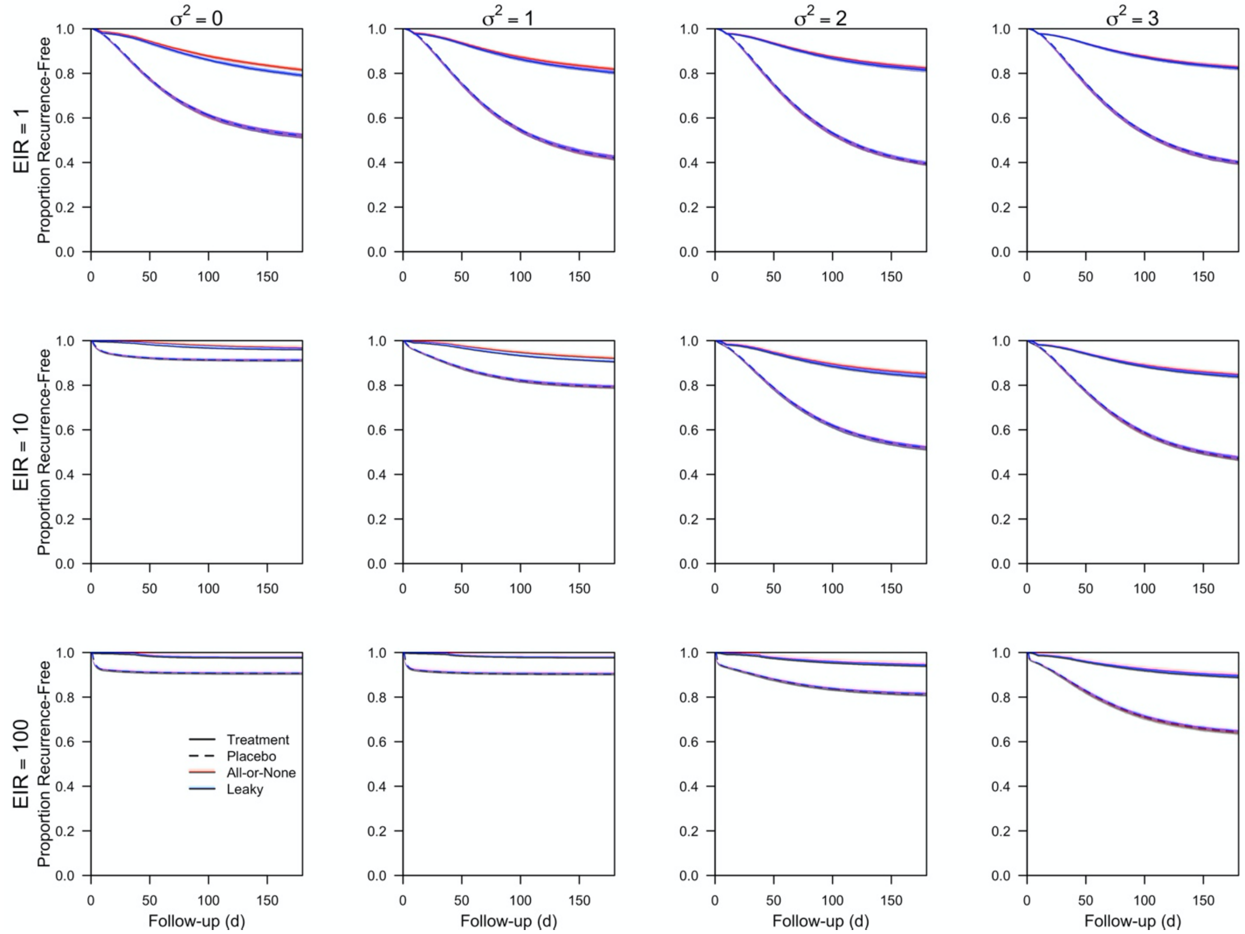


***Figure S10. Recurrence-free survival curves for "all-or-none" and "leaky" intervention actions.*** *Recurrence-free survival curves from simulated clinical trials as a function of follow-up time are shown at different entomological inoculation rates (EIR) and heterogeneity in biting ($\sigma^2$) for "all-or-none" (red) and "leaky" (blue) intervention actions. The infection endpoint used is all LM-detectable recurrent infections. Each line is the median of 200 simulations, and each region is the interquartile range. Solid lines correspond to the treatment arm, and dashed lines correspond to the placebo arm.*

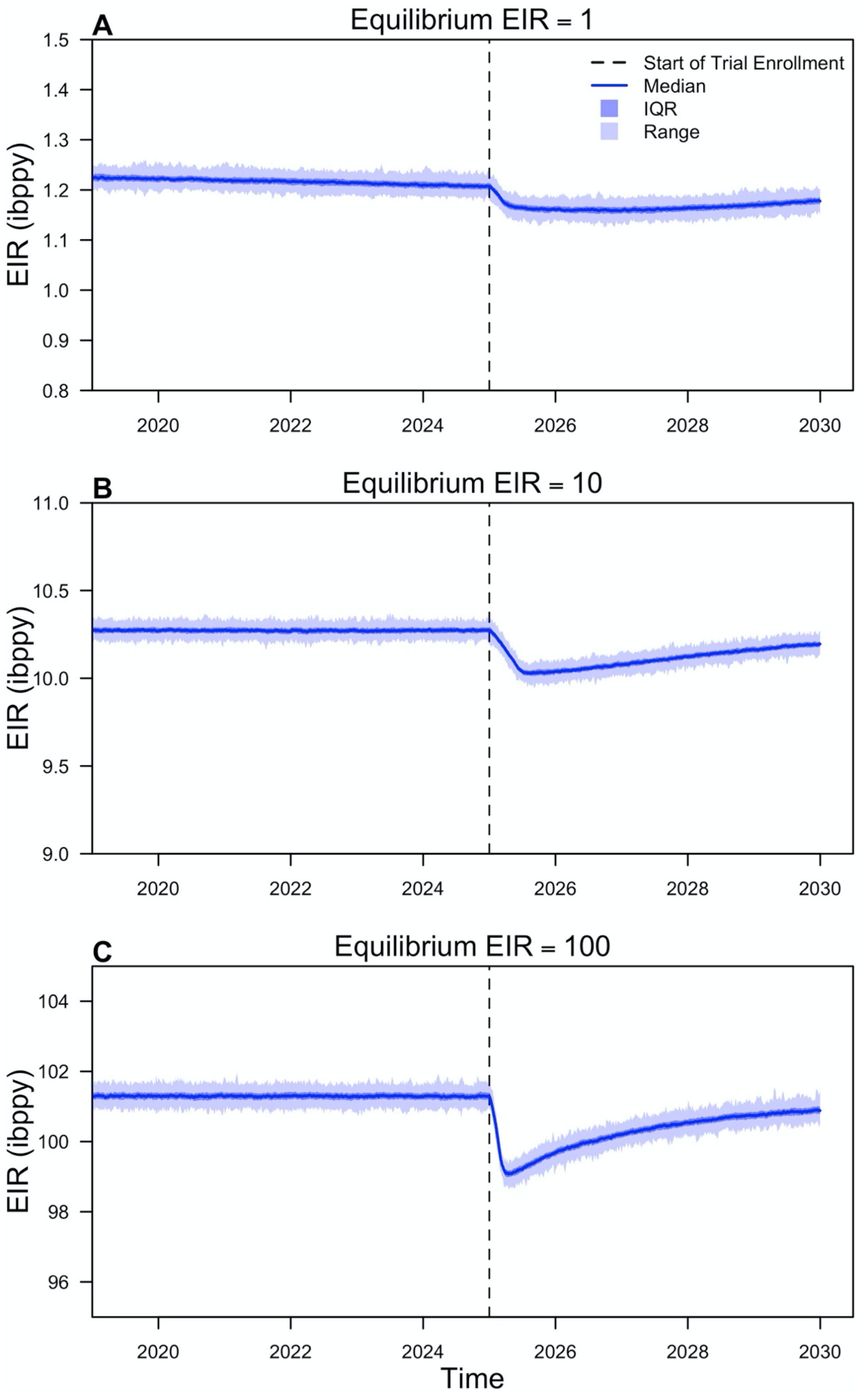

A
Equilibrium EIR = 1
Start of Trial Enrollment
Median
IQR
Range
EIR (ibppy)
1.5
1.4
1.3
1.2
1.1
1.0
0.9
0.8
2020
2022
2024
2026
2028
2030
B
Equilibrium EIR = 10
EIR (ibppy)
11.0
10.5
10.0
9.5
9.0
2020
2022
2024
2026
2028
2030
C
Equilibrium EIR = 100
EIR (ibppy)
104
102
100
98
96
2020
2022
2024
2026
2028
2030
Time


487

***Figure S11. Entomological inoculation rate over time.*** *The entomological inoculation rate (EIR) experienced by the population is shown over the full simulation period when the equilibrium EIR was (A) 1, (B) 10, and (C) 100. The blue line denotes the median, the darker shaded region denotes the interquartile range, and lighter shaded region denotes the full range. The dotted black line indicates the start of each trial.*
